# Spatially-resolved wastewater-based surveillance enables COVID-19 case localization across a university campus, and confirms lower SARS-CoV-2 RNA burden relative to the surrounding community

**DOI:** 10.1101/2023.03.03.23286756

**Authors:** Jangwoo Lee, Nicole Acosta, Barbara J. Waddell, Kristine Du, Kevin Xiang, Jennifer Van Doorn, Kashtin Low, Maria A. Bautista, Janine McCalder, Xiaotian Dai, Xuewen Lu, Thierry Chekouo, Puja Pradhan, Navid Sedaghat, Chloe Papparis, Alexander Buchner Beaudet, Jianwei Chen, Leslie Chan, Laura Vivas, Paul Westlund, Srijak Bhatnagar, September Stefani, Gail Visser, Jason Cabaj, Gopal Achari, Rhonda G. Clark, Steve E. Hrudey, Bonita E. Lee, Xiaoli Pang, Brandan Webster, William Amin Ghali, Andre Gerald Buret, Tyler Williamson, Danielle A. Southern, Jon Meddings, Kevin Frankowski, Casey R.J. Hubert, Michael D. Parkins

## Abstract

Wastewater-based surveillance (WBS) has been established as a powerful tool that can guide health policy at multiple levels of government. However, this technology has not been well assessed at more granular scales, including large work sites such as University campuses. Between August 2021-April 2022, we explored the occurrence of SARS-CoV-2 RNA in wastewater from multiple complimentary sewer catchments and residential buildings spanning the University of Calgary’s campus and how this compared to levels from the municipal wastewater treatment plant servicing the campus. Concentrations of wastewater SARS-CoV-2 N1 and N2 RNA varied significantly across six sampling sites – regardless of several normalization strategies – with certain catchments consistently demonstrating values 1–2 orders higher than the others. Additionally, our comprehensive monitoring strategy enabled an estimation of the total burden of SARS-CoV-2 for the campus per capita, which was significantly lower than the surrounding community (p≤0.01). Real-time contact tracing data was used to confirm an association between wastewater SARS-CoV-2 burden and clinically confirmed cases proving the potential of WBS as a tool for disease monitoring across worksites. Allele-specific qPCR assays confirmed that variants across campus were representative of the community at large, and at no time did emerging variants first debut on campus. This study demonstrates how WBS can be efficiently applied to locate hotspots of disease activity at a very granular scale, and predict disease burden across large, complex worksites.

**Synopsis:** ‘This study establishes that wastewater-based surveillance with a node-based sampling strategy can be used to passively monitor for disease, locate disease “hotspots” and approximate the burden of infected individuals’

## 1. Introduction

To cope with the COVID-19 pandemic crisis, governments worldwide have implemented a range of measures to mitigate the spread of the virus, including large-scale clinical testing. While diagnostic testing is tremendously important, it is limited in its capacity to perform population-level surveillance owing to tremendous human and capital resources to run community testing centers.^1^ In addition, clinical testing is biased by relying on voluntary participation and towards individuals with symptomatic disease.^2^ Wastewater-based surveillance (WBS) has emerged as a novel tool for monitoring population health^3^ serving to complement clinical testing providing real-time data on the burden of disease in a monitored sewershed. Advantages of WBS include i) comprehensive, inclusive, serial monitoring of the population with relatively low costs, and ii) unbiased data collection of biological material from all society members including marginalized populations and those unable to access clinical testing.^1, 3, 4^ WBS for SARS-CoV-2 surveillance has been adopted worldwide^5^, including Canada where it currently covers ∼62 % of the country’s population.^6^ Generally, WBS programs monitor SARS-CoV-2 RNA in untreated sewage from wastewater treatment plants (WWTPs) and thus assess disease burden at the level of an entire community. Community-based studies have identified SARS-CoV-2 WBS as a leading indicator for cases (4-6 days), hospitalizations, and deaths.^7-10^ However, studies where monitoring has been performed at a more granular scale (e.g., defined sub-catchments within a larger sewershed or specific facilities) are less common, and more work is needed to clearly demonstrate the similar benefits in more local contexts with sewer catchments that enable wastewater sampling corresponding to smaller zones.

Such granular scale WBS is challenging when it comes to selecting sampling locations because i) it requires detailed data on defined sampling nodes (e.g., each node and their GPS coordinates) and their connectivity (to determine if overlapping catchments exist and may confound analysis), ii) nodes should have enough wastewater flow to ensure sufficient volume during continuous collection. A recent study in the City of Calgary evaluating neighborhood-scale sub-catchment monitoring (serving populations of 13,000 to 73,000) within larger WWTP catchments (serving 290,000 to 1,048,000) showcased the utility of node-based sampling in identifying specific sub-catchment(s) with disproportionate infection burden.^11^ WBS at an even more granular scale (e.g., buildings) as a node-based sampling strategy could help to specify more clearly ‘hotspots’ for infection transmission.^12, 13^

One area of focused WBS that has generated significant interest is university/college campuses. University campuses consist of a vast array of building complexes where a high degree of social interaction is expected between students, staff, and faculty. High rates of SARS-CoV-2 transmission are possible in the absence of mitigating steps. While there have been university campus-wide studies, spatial distribution of SARS-CoV-2 RNA within the university campus, and studies enabling a comprehensive assessment of the total disease burden have not as yet been performed. Thus far, University-based studies assessing SARS-CoV-2 RNA in wastewater have primarily focused on longitudinal analysis rather than spatially resolved analysis^14, 15^ or only on individual dormitories/residential buildings^16-18^, limiting their impact.

Herein we describe a longitudinal nodal-based WBS monitoring program for SARS-CoV-2 RNA at the University of Calgary (UofC), Calgary, Alberta, Canada. The main campus is situated on 530-acres in the Northwest quadrant of the city and were monitored using WBS from Aug 31, 2021–Apr 24, 2022, along with the municipal WWTP which serves the campus. Our primary research objectives were to i) locate specific sub-catchment(s) within the University where SARS-CoV-2 RNA exists in differential abundance and associate this with COVID-19 case occurrence, and ii) determine the relative risk of COVID-19 on campus, as inferred by SARS-CoV-2 wastewater burden, relative to the surrounding community. We hypothesized that i) higher abundance of SARS-CoV-2 RNA would be found in buildings with higher social connectivity and that this would be associated with COVID-19 reported cases; and, ii) the abundance of SARS-CoV-2 RNA in campus wastewater would be lower than the surrounding community given a highly educated population, campus mandate for universal masking, a high prevalence of accessible hand hygiene product through campus, and a vaccine mandate (or weekly negative test) required to attend campus.

## 2. Materials and Methods

### 2.1. Defining sampling nodes across the UofC sewershed

UofC is among the ten largest Universities in Canada with >26,000 full-time undergraduate students, 6,000 graduate students and 1,800 academic and 3,200 non-academic staff.^19^ The main campus is situated in Northwest Calgary on a parcel of 530-acres (2.13 km^2^). Campus sampling locations were chosen to deliver a minimum number of manhole-accessible sites providing maximum coverage of campus buildings, using GIS-based analysis of the sewer pipe network (Fig. 1). Residence halls were also included based upon presence of an accessible sampling location within the building’s plumbing network capturing ≥50% of the residential areas of that building. This was accomplished by manually reviewing the engineering drawings of the dormitory plumbing systems, visually inspecting each prospective sampling location and then selecting the site that optimally captured the building’s wastewater in a safe, access-controlled manner.

**Figure 1.**
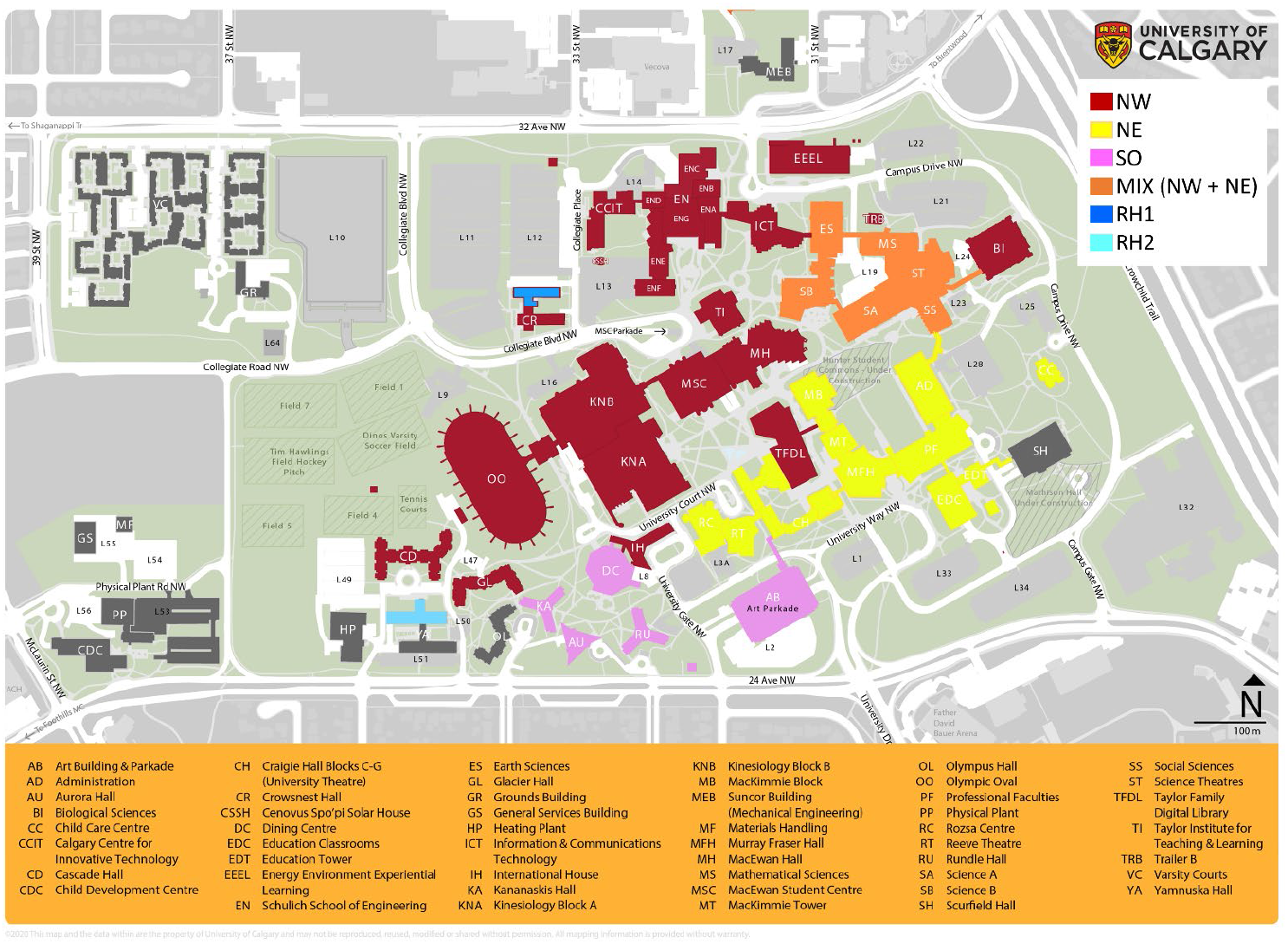
University of Calgary main campus highlighting different sewershed catchments (colour key is shown in the figure legend). NW, NE, and SO indicate Northwest, Northeast, and South catchments, respectively. RH1 and 2 represent the two student residence halls. The area MIX (colored in orange) belongs to both NW, and also NE. Five university buildings outside our monitoring catchments, but which still belong to the main campus were colored in ‘dark grey’. L1 – 64 (in light grey) represent un-serviced parking lots. The figure was modified from http://www.ucalgary.ca/map.

Three nodes were selected to capture the building complexes in the Northwest (NW), Northeast (NE), and South (SO) zones of the campus (Fig. 1). Six buildings drain into both NW and NE within these catchments and are indicated as MIX (see Fig. 1; colored in orange). Both residence halls (RH1 and RH2) are within the NW catchment thus enabling even more granular scale sampling nodes. The entire monitoring program captured 7 residential halls, 39 lecture/research facilities and 5 recreational facilities (including dining/fitness buildings) (Fig. 1). Our monitoring catchments cover >80% of the total residence population (i.e., those living in dormitories) and >83 % of the campus aerial footprint.

Calgary is Canada’s fourth largest city by population and its third most ethnically diverse.^11, 20^ Three WWTPs serve an estimated 1,441,268 people.^21^ UofC falls exclusively within the catchment zone of the largest WWTP, serving 1,047,662 individuals and receiving 303.7-604.6 ML/day.

### 2.2. Wastewater Collection

Wastewater samples were collected from the sites described above 2-3× per week from August 31, 2021–April 30, 2022, using a workflow described previously.^11, 12^ In short, CEC Analytics V1 (C.E.C Analytics, Canada) and ISCO 6712 (Teledyne ISCO, USA) autosamplers collected 2L (CEC) or 10L (ISCO) 24-hours composite samples that were stored at 4°C and transported to Advancing Canadian Water Assets (ACWA). Upon arrival, samples were thoroughly mixed and aliquoted into 50mL centrifuge tubes for downstream analysis. More details on field sampling techniques are described in the Supplement.

### 2.3. Sample processing and RNA extraction

Sample processing and RNA extraction was performed following established workflows.^11, 12, 22, 23^ In brief, 40mL of thoroughly mixed wastewater subsamples was added to 50mL falcon tubes prefilled with 9.5 g of sterile NaCl and 400ul of TE buffer and were then spiked with 200µL of Bovine Coronavirus (BCoV) (final concentration: 5×10^5^ 50% tissue-culture-infective dose (TCID_50_) per mL) as an internal control and vortexed for 30 seconds. Solids were then removed via vacuum filtration through a 5 µm polyvinylidene difluoride membrane, where samples were filtered directly into 40ml of 70% ethanol. This solution was then passed through a Zymo Spin™ III-P silica column (Zymo Research, USA). ^24^ More details can be found in the Supplement.

### 2.4. Quantitative RT-qPCR

RT-qPCR assays were performed following established workflows.^11, 12, 24^ In short, two regions of the nucleocapsid gene (N1 and N2) were used to quantify total SARS-CoV-2 RNA copies/mL in every wastewater sample.^25-27^ We also analyzed variants of concern (VOC), including Delta, Omicron (BA.1, and BA.2) using the N200 multiplex assay^24,28^ or 69/70del assay^29^ for a subset of samples for WWTP (44 samples), RH1 (18 samples), SO (18 samples), NW (17 samples), and NE (9 samples) from November 28, 2021-April 27, 2022. BCoV (Bovine Coronavirus) was analyzed as an internal spike control, and PMMoV (Pepper Mild Mottle Virus) was analyzed as a potential human feces biomarker.^12, 23, 30^ All samples were analyzed in triplicate, including non-template controls for each run using QuantStudio-5 Real-Time PCR System (Applied Biosystems, USA). Samples with a quantification cycle (Cq)<40 were considered positive.^11, 12^ Key quality parameters (i.e., efficiency, R^2^ of regression curve, Y-intercept, and slope) for qPCR standard curves are shown in Table S1. Detailed information (e.g., oligonucleotide sequences and thermal cycling conditions) can be found in previous publications.^11, 12, 24^ The raw data were subjected to further quality control and performed similarly to previously published works(see Supplementary Results 2.1).^11, 12, 24^ Finally, concentrations for N1, N2, and PMMoV were averaged weekly for each monitoring location. Those averaged concentrations (for each week) were used in most downstream analyses, and the data presented in Dataset S1.

### 2.5. Chemical analysis

In addition to PMMoV, a total of five wastewater cations (sodium, chloride, potassium, magnesium, and calcium) were chosen to explore their association with SARS-CoV-2 as potential normalization markers for human activity. These cations are associated with human excreta, especially urine^31, 32^ and potentially useful for correcting possible underestimation of SARS-CoV-2 levels due to dilution effect^33, 34^, which could be particularly important in small catchments. Relationships between N1 and N2 gene abundance and wastewater cations were examined for samples collected during August 31, 2021 to January 04, 2022 (n=13 to 36 depending on sampling site). To analyze cations, wastewater samples were thoroughly homogenized, and filtered through 1.5μm dried pre-rinsed Grade 934-AH® RTU glass microfiber filters (Whatman, UK) by 12mL. The filtrate was filtered again through a 0.45μm PVDF membrane UNIFLO® syringe filter (Whatman, UK), distributed to a tube for ion chromatography, and then stored at 4ºC until analysis. Those processed samples were analyzed using Metrohm 930 Compact Ion Chromatography Flex (Metrohm, Switzerland).

### 2.6. Modelling expected COVID-19 cases per capita across UofC Campus

Cases per capita in UofC main campus (*CPC*_*Uofc*_) was calculated using raw (i.e. un-normalized) SARS-CoV-2 RNA concentrations according to the following relationship referring to (Eq.S4) in the Supplement

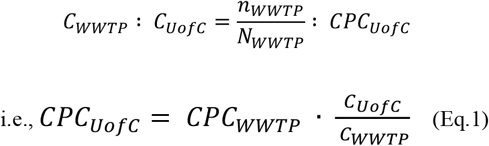

Where, *N*_*WWTP*_ indicates the total population in the catchment area for WWTP (n=1,047,622^11^ ; *n*_*WWTP*_is incident number of new cases occurring daily (i.e., confirmed COVID-19 infected individuals) in the catchment area for WWTP; *C*_*Uofc*_ and *C*_*WWTP*_indicate concentration of SARS-CoV2 RNA for UofC and WWTP respectively, and could be calculated according to (Eq.S6) in the Supplement.

To mitigate uncertainty which may arise from possible differences in human excreta across samples, *CPC*_*Uofc*_ was also calculated using normalized SARS-CoV-2 RNA concentration according to the following relationship referring to (Eq.S5) in the Supplement.

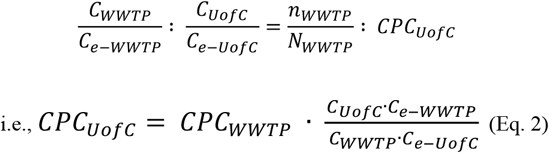

Where, *C*_*e*−*Uofc*_, and *C*_*e*−*WWTP*_indicate concentration of human excreta surrogates for UofC and WWTP, and could be calculated according to (Eq. S6) in the Supplement.

### 2.7. Uncertainty analysis

As wastewater flow data for UofC was unavailable, models described in 2.6 rely on assuming that flow quantities are proportional to catchment surface areas (i.e., total footprint of all buildings) (see Supplementary Method 1.3 in the Supplement). We assumed that uncertainty in this model derives mostly from variability of gross surface area in prediction of flow quantity. Therefore, prediction errors from these sources were propagated using a Monte-Carlo randomization simulation adapted from other relevant works.^35, 36^ The term surface area (*A*) was randomized by multiplying the uncertainty factor (*a*) which is variable ranging from 0.2 (20%) to 2.0 (200%) assuming that the actual ratio of flow quantity lies within these boundaries.

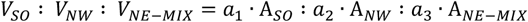

Where, *a*_1_, *a*_2_, and *a*_3_ are random variables ranging from 0.1 to 2.0, also are independent from each other.

The simulation was repeated 1,000 times, and interquartile ranges (IQR, Q1 – Q3) for each prediction value are displayed as error bars in the model. Furthermore, the p-value for permutation test was defined as ‘the ratio of counts where *CPC*_*Uofc*_>*CPC*_*WWTP*_ to 1,000 (= the number of simulation trials)’ for each time point. Only time-paired data points were compared between each site. The modelling was performed using R (v4.1.2), and related datasets/R codes will be available in the first author’s GitHub page (https://github.com/myjackson) upon acceptance of this manuscript.

### 2.8. Clinical case documentation

Information on city-wide, new daily cases of clinically confirmed COVID-19 (patient swabs confirmed with a clinical RT-qPCR) were provided by a single comprehensive public health system, Alberta Health Services (AHS) via the Centre for Health Informatics online COVID Tracker (https://covid-tracker.chi-csm.ca/). The information was gathered between August 31, 2022, and March 31, 2022, and a subset of this data (i.e., August 31, 2021 – January 04) was subjected to further analysis. New cases were binned by individual postal codes (using the first three of six digits) as an indicator of the home address of newly diagnosed cases. Cases were then assigned to the appropriate WWTP serving their primary residence.

Comparative analyses were conducted across two distinct time periods; Period A (Aug 31, 2021-Dec 12, 2021) and Period B (Dec 13, 2021-April 25, 2021) owing to fundamental changes occurring through the pandemic. In particular, during the Omicron waves (Period B, defined when the first case was documented in Calgary), clinical case occurrence for the first time vastly exceeded the ability of health services to screen and detect the population.

Documenting campus-associated confirmed-COVID-19 cases and ascribing them to a specific primary building was performed in real-time by the University of Calgary’s Occupational Health and Staff Wellness for students and employees who self-reported a positive COVID-19 test during the pandemic period from September 2021. Confirmed cases were excluded from attending campus for a minimum of 10 days (reduced to 5 days after January 3^rd^, 2022) and complete symptom resolution. The information gathered between September 21, 2021–April 2022 was used to trace the primary buildings where individuals with confirmed COVID-19 were located. We assigned study-specific personal identifiers to each affected individual and avoided personal identifying information. The information gathered in the original report includes: i) ‘date of positive test result’, ii) ‘date university informed of illness’, iii) ‘recent university location(s) visited and the date when the person visited that location’, and iv) ‘date of onset of symptoms’, etc. However, in some instances case information was not always fully declared (i.e., the recent university location(s) visited, and the date(s) when the person visited), and such cases were excluded. As a result, the information from 463 out of 721 reported individuals was used in downstream analyses. The raw data is not shown for ethical consideration, but could be provided upon reasonable request to the authors. The full set of processed data is shown in Dataset S2. The patient identifiers (PID) are not known to anyone outside our research group, so individuals remain anonymous.

To model the movement of confirmed COVID-19 infected individuals across campus, we relied on self-reported activity tracing. To identify individual buildings where COVID-19 positive individuals visited, we first counted the number of positive individuals who visited each building using the information iii) above. For example, for each building, the recently visited PIDs were listed (Dataset S2). Then, we counted the total affected-visits for each building. In this way, each PID was often counted multiple times in situations where the person visited multiple locations or one location on multiple days during the monitoring period. As the majority of SARS-CoV-2 RNA shedding occurs in the few days before and after symptom onset^12, 22^, the visits ± 2 days of the ‘date of onset of symptoms’ were considered valid, otherwise excluded in the following analysis. Total affected-visits is named ‘number of cases’. Finally, the number of cases was subjected to further analysis. For instance, the cases were averaged weekly, and aggregated by monitoring catchment for each monitoring week (Fig. S5).

During the monitoring period, on-campus residents (i.e. those residing full-time in dormitories) who were confirmed as COVID-19 positive were quarantined according to the following principles: i) if a case was reported by an individual living in a single unit with a bathroom– not shared with another, the individual was isolated in place (a total of 10 residential halls, CR, YA, CD, GL, KA, AU, RU, IH, OL, or VC (Fig. 1)), ii) if all occupants of a shared apartment are positive, they would continue to isolate in their same apartment in their residential hall, and iii) if the positive individual shares an apartment with someone who is not also positive, they were moved to another suite, VC for their isolation period. Our monitoring program included most of the isolation places (i.e., a total of 8 out of 10 places, CR, YA, CD, GL, KA, AU, RU, and IH; see Fig. 1).

### 2.9. Statistical analysis

Kruskal-Wallis test followed by a post-hoc Wilcoxon rank-sum test was performed to test if there were significant differences between groups. For pairwise tests, p-values were adjusted using the Benjamini-Hochberg method. Additionally, Spearman correlation analysis was performed to test if there were significant relationships between the two factors. Finally, Fisher’s exact test was implemented to test the potential association between two variables (i.e., SARS-CoV-2 signals versus campus-associated COVID-19 cases). One-sided test was employed under the expectation that those two might be positively associated. Then, Fisher’s exact test was repeated for each pair of wastewater SARS-CoV-2 signals (N1 or N2) against cases reported; a week earlier (−1 week) cases, those on the same week (+0 week), or the week following (+1 week) under the hypothesis that wastewater signals serve as an early warning sign of COVID-19 cases. The key rationale was explained in more detail in the Supplementary Method 1.5. All analyses were done using R version 4.1.2., and related datasets/R codes will be available in the first author’s GitHub page (https://github.com/myjackson) upon acceptance of this manuscript.

### 2.10. Ethics

This study received approval from the Conjoint Research Health Ethics Board of the University of Calgary (REB20–1544).

## 3. Results

### 3.1. Longitudinal tracking wastewater-borne SARS-CoV-2 RNA across campus

Between August 3, 2021–April 30, 2022, a total of 58 (RH1) and 18 (RH2) samples were obtained from the residence halls, 45 (NE), 48 (SO), and 42 (NW) samples were obtained from the campus catchments, and 89 samples were obtained from WWTP, providing 12 – 25 data points per location after being averaged weekly. The lower number of samples collected for campus locations primarily relates to more complicated access for these sampling points (e.g., manholes in the open (public) area for the campus sampling points versus either within buildings or from the WWTP facility. The outdoor locations (manholes) also experienced a higher rate of failure to collect especially during cold weather (<-20 ?C) for campus sites.

During this time the City of Calgary experienced three successive “waves” of COVID-19 (corresponding to the fourth, fifth and sixth waves since the start of the pandemic). Tracked via wastewater, the first of these waves during the monitoring period peaked on September 06, 2021, followed by January 03 and April 18, 2022. Allele-specific PCR to detect VOC in WWTP samples confirmed it was the Delta variant that was dominant during the fourth-wave (peaking September 06, 2021), and Omicron lineages were dominant during the fifth (BA.1 peaking January 03, 2022) and sixth waves of this study (BA.2 peaking April 18, 2022, the third wave in this study) (Fig. S6). Notably, the burden of wastewater-detected SARS-CoV-2 N1 and N2 for the two latter waves caused by Omicron lineages vastly exceeded that of Delta.

SARS-CoV-2 N1 and N2 concentrations across campus monitoring sites generally, but not always mirrored those of the community WWTP (Fig. 2). Values from WWTP were higher than those across campus, with some exceptions. While the highest N1 and N2 values observed from campus monitoring sites (i.e., SO, NE, and NW) occurred during the peaks of each wave experienced in the community, random spikes in N1 and N2 also occurred during community troughs and appeared randomly suggesting brief periods of increased disease burden. Analysis of VOC across UofC campus mirrored those for WWTP –Delta was dominant in Period-A (Aug 31, 2021-Dec 12, 2021) for those locations where data was available (i.e., SO, and NW) (Fig. S7 & S8) and Omicron lineages were dominant in Period-B for SO, NW, NE, and RH1 while. In no instances did the emerging VOC occur disproportionally within the campus environment relative to that of the community.

**Figure 2.**
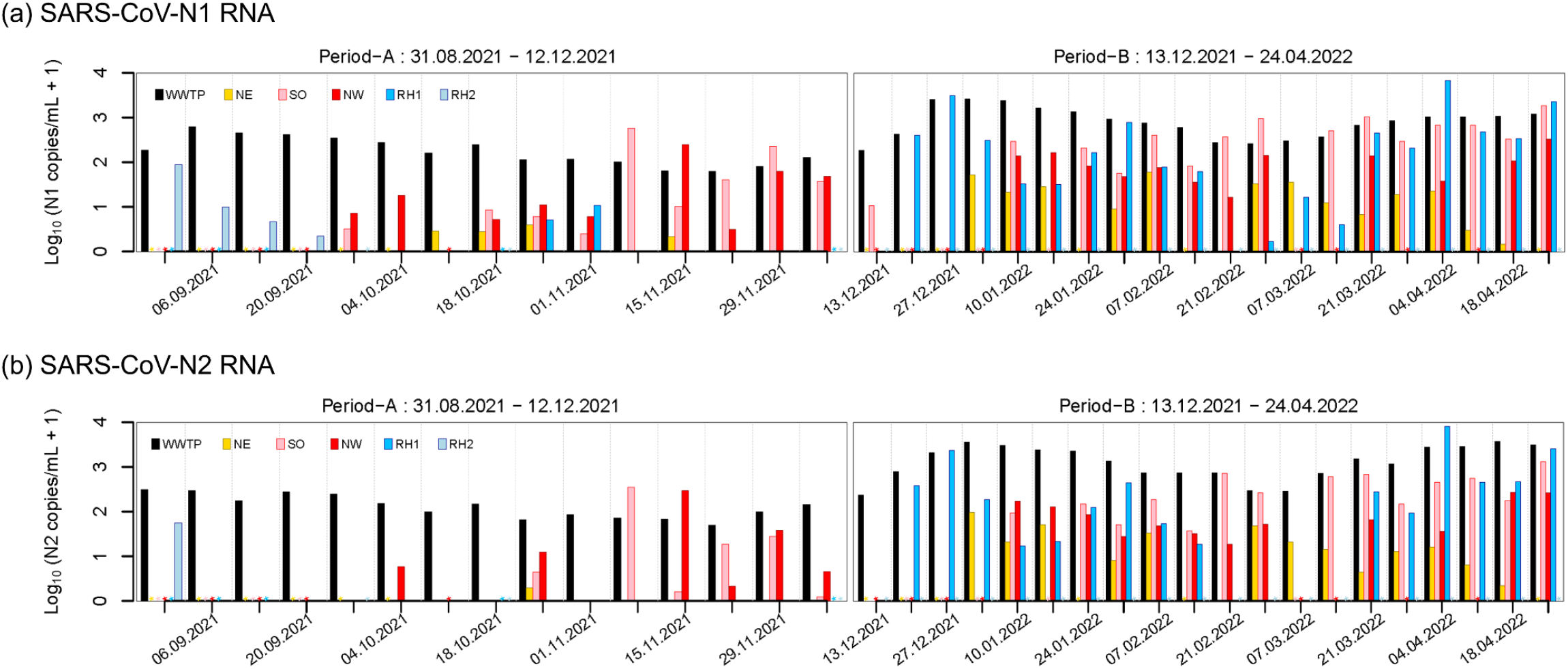
Log_10_-transformed concentrations (copies/mL) of SARS-CoV-2 N1 and N2 profiles in campus wastewater sub-catchments demonstrate much lower values relative to the receiving municipal WWTP during the study period (August 31, 2021 – April 30, 2022). WWTP indicates the municipal wastewater treatment plant servicing the surrounding community, and also UofC main campus. See Fig. 1 for the locations and catchment area. *=missing data. Date (in x-axis) : dd.mm.yyyy.

### 3.2. Correlating wastewater SARS-CoV-2 RNA with clinically confirmed cases

A median of 153 (IQR 73 – 240) cases per day were clinically confirmed across the catchment of the WWTP during the period monitored (August 31, 2021 – January 04, 2022; a total of 36 data points). These cases were correlated with raw-, and also normalized-SARS-CoV-2 N1 and N2 signals using different investigational markers (i.e., PMMoV, sodium, chloride, potassium, calcium, and magnesium) for the corresponding date ranges. The raw (i.e., un-normalized) N1 and N2 signals (i.e., copies/mL) correlated with confirmed cases the best (where N2 was more sensitive than N1) suggesting biomarkers for normalization did not denoise variability associated with human excreta over time. However, for comparing different smaller catchment results with each other, we expected the variability in human wastes between sites could be large, especially when the characteristics of those populations may be very different, e.g., residential versus non-residential areas of the campus. Accordingly, while we used raw SARS-CoV-2 concentrations as our primary outcome for intra-site comparisons, PMMoV and ion normalization were assessed as a confirmatory secondary outcome measure.

### 3.3. Comparing SARS-CoV-2 RNA signals across different sampling locations

Comparison across different locations was conducted for each of the two separate periods, for instance Period-A (Aug 31, 2021-Dec 12, 2021) and B (Dec 13, 2021-April 25, 2021). There were significant differences in both raw and normalized wastewater SARS-CoV-2 RNA concentrations between monitoring sites during the study (Period-A to -B) based on Kruskal-Wallis test (p<0.001). In Period-A, a post-hoc analysis using Wilcoxon-rank sum test revealed that SARS-CoV-2 RNA N1 and N2 concentrations in campus locations were 1 – 2 orders of magnitude lower than the community WWTP (p≤0.005) (Table 1 & Fig. 3). Furthermore, there were significant differences in both N1 and N2 concentrations between campus locations. For instance, the values for NE were 1 – 2 orders of magnitude lower than NW and SO, and such difference was non-parametrically significant using Wilcoxon rank-sum tests for NW (p≤0.038) (Table 1). SARS-CoV-2 N1 and N2 concentrations for two dormitories (i.e., RH2 and RH1) were similar to NE (p≥0.428) but lower than SO (p≤0.026) and NW (p≤0.038), based on Wilcoxon rank-sum test (Table 1). In general, normalized trends of SARS-CoV-2 burden between sites mirrored those of raw values (Table S3). The normalized N1 and N2 concentrations for WWTP were higher than for all the campus locations using post-hoc Wilcoxon rank-sum test in all comparisons (p≤0.008) (Table S3). Among university campus locations, the normalized N1 and N2 values for NE were lower than NW or SO in many instances (Table S3).

**Table 1.** Comparing SARS-CoV-2 RNA raw concentrations between different UofC monitoring locations during Period-A and -B using Wilcoxon rank-sum test. P-adjusted for pairwise comparison using Benjamini & Hochberg method. Those pairs that statistically differed with (p<0.05) are highlighted in red.

| Period | Indicator | Location | RH2 | RH1 | NE | SO | NW |
| --- | --- | --- | --- | --- | --- | --- | --- |
| Period-A | N1 | RH1 | 0.675 | - | - | - | - |
|  |  | NE | 0.968 | 0.650 | - | - | - |
|  |  | SO | 0.026 | 0.014 | 0.020 | - | - |
|  |  | NW | 0.014 | 0.005 | 0.005 | 0.863 | - |
|  |  | WWTP | 0.000 | 0.000 | 0.000 | 0.005 | 0.001 |
|  |  | NE | 0.944 | 0.428 | - | - | - |
|  |  | SO | 0.052 | 0.020 | 0.068 | - | - |
|  |  | NW | 0.038 | 0.015 | 0.038 | 0.736 | - |
|  |  | WWTP | 0.000 | 0.000 | 0.000 | 0.002 | 0.002 |
| Period-B | N1 | RH1 | - | - | - | - | - |
|  |  | NE | - | 0.019 | - | - | - |
|  |  | SO | - | 0.156 | 0.000 | - | - |
|  |  | NW | - | 0.491 | 0.002 | 0.006 | - |
|  |  | WWTP | - | 0.003 | 0.000 | 0.013 | 0.000 |
|  |  | NE | - | 0.107 | - | - | - |
|  |  | SO | - | 0.238 | 0.001 | - | - |
|  |  | NW | - | 0.829 | 0.006 | 0.029 | - |
|  |  | WWTP | - | 0.000 | 0.000 | 0.000 | 0.000 |

**Figure 3.**
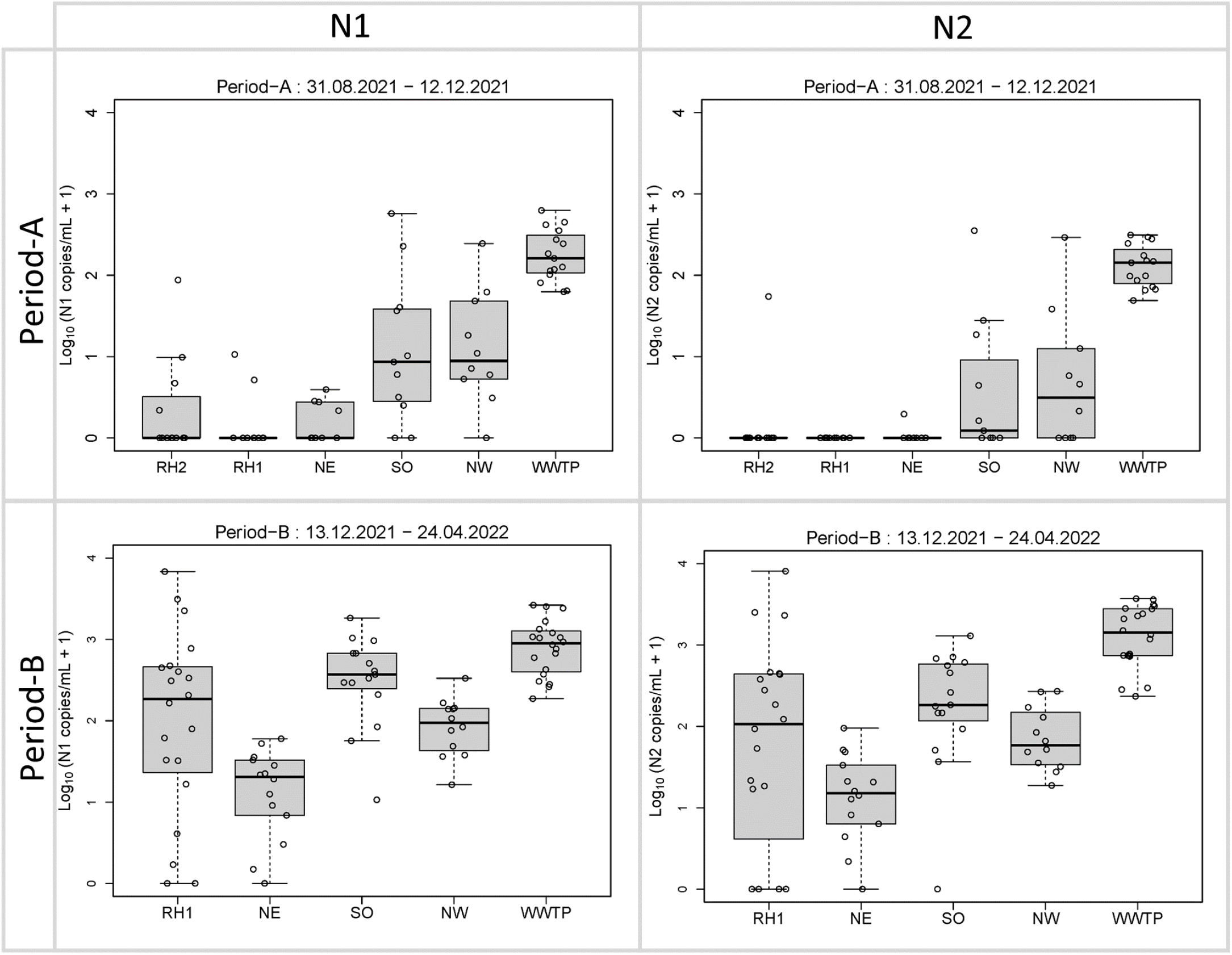
Log_10_-transformed concentrations of SARS-CoV-2 N1 (left) and N2 (right) by sub-catchment monitoring location variably demonstrate differences between locations during Period-A (Aug 31, 2021-X) and -B (Y-April 30, 2022) with (upper), and without normalization (lower). See Table 2 and 3 for the results from Wilcoxon rank-sum tests.

In Period-B, the N1 and N2 concentrations increased considerably compared to Period-A at WWTP and across campus (Fig. 3). However, the values for the campus were still significantly lower than for WWTP for both N1 and N2 based on a post-hoc Wilcoxon rank-sum test (p≤0.013; see Table 1). The degree of increase for RH1 was the most pronounced among all monitored sites. For instance, the median N1 concentration for RH1, and SO samples profoundly increased, for instance by >2 order of magnitude (from 0.0 to 184.7 copies/mL for RH1; from 7.6 to 369.8 copies/mL), while the median concentration for other campus locations increased only by approximately 1 order of magnitude (from 0.0 to 19.4 copies/mL for NE; from 8.1 to 94.3 copies/mL for NW). In all cases, the normalized concentrations for SO were significantly higher than for NE based on Wilcoxon rank-sum tests in all cases using N1 (p≤0.025), and most cases for N2. The normalized concentrations for WWTP were significantly higher than all the UofC campus locations in all cases using N2 (p≤0.014), and in many cases except for chloride, sodium, and potassium using N1 (p≤0.036) (Table S4).

**Table 2.** The Fisher’s exact test results (p-value) for testing interdependency between wastewater SARS-CoV-2 signals and COVID-19 confirmed cases across UofC campus under each assumption. ‘-1 week’, ‘+0 week’, and ‘+1 week’ indicate an early warning, no time lag, and time lag scenario, respectively (see Fig. S1 in the Supporting Information for details). The results with p < 0.05 were highlighted in red.

| Location | N1 |  |  | N2 |  |  |
| --- | --- | --- | --- | --- | --- | --- |
|  | (-1 week) | (+0 week) | (+1 week) | (-1 week) | (+0 week) | (+1 week) |
| RH1 | 0.001 | 0.000 | 1.000 | 0.001 | 0.000 | 1.000 |
| RH2 | 0.273 | 0.333 | 0.333 | 1.000 | 1.000 | 0.083 |
| NE | 0.029 | 0.008 | 0.154 | 0.029 | 0.071 | 0.433 |
| SO | 0.021 | 0.205 | 0.163 | 0.002 | 0.048 | 0.163 |
| NW | 0.099 | 0.193 | 0.063 | 0.099 | 0.193 | 0.063 |

**Table 3.** Comparing modelled COVID-19 cases per capita between WWTP and UofC campus using raw and normalized values. The p-values were calculated using Permutation test (see 2.7), and indicate the proportion of the number of cases where cases per capita for UofC > cases per capita for WWTP to 1,000 (=the number of total simulation runs). Those pairs that statistically differed with (p < 0.05) are highlighted in red.

| Date | Raw | Normalized |  |  |  |  |  |
| --- | --- | --- | --- | --- | --- | --- | --- |
|  |  | PMMoV | Magnesium | Potassium | Sodium | Calcium | Chloride |
| 18.10.2021 | <0.001 | 0.003 | 0.003 | 0.003 | 0.003 | 0.003 | 0.003 |
| 25.10.2021 | <0.001 | 0.003 | 0.003 | 0.003 | 0.003 | 0.003 | 0.003 |
| 01.11.2021 | <0.001 | 0.003 | 0.003 | 0.003 | 0.003 | 0.003 | 0.003 |
| 08.11.2021 | 0.247 | 0.278 | 0.328 | 0.578 | 0.809 | 0.318 | 0.658 |
| 15.11.2021 | 0.981 | 0.972 | 0.999 | 0.999 | 0.999 | 0.999 | 0.999 |
| 22.11.2021 | <0.001 | 0.003 | 0.003 | 0.003 | 0.003 | 0.003 | 0.003 |
| 29.11.2021 | <0.001 | 0.003 | 0.003 | 0.003 | 0.007 | 0.003 | 0.003 |
| 06.12.2021 | <0.001 | 0.003 | 0.003 | 0.003 | 0.003 | 0.003 | 0.003 |
| 10.01.2022 | <0.001 | 0.003 | 0.003 | 0.003 | 0.003 | 0.003 | 0.003 |
| 31.01.2022 | <0.001 | 0.003 | 0.003 | 0.003 | 0.003 | 0.003 | 0.003 |
| 07.02.2022 | <0.001 | 0.003 | 0.003 | 0.003 | 0.003 | 0.003 | 0.003 |
| 21.02.2022 | <0.001 | 0.003 | 0.003 | 0.003 | 0.022 | 0.003 | 0.003 |
| 28.02.2022 | <0.001 | 0.003 | 0.003 | 0.003 | 0.124 | 0.003 | 0.004 |
| 21.03.2022 | <0.001 | 0.003 | 0.003 | 0.003 | 0.003 | 0.003 | 0.003 |

### 3.4. SARS-CoV-2 RNA measured in campus wastewater catchments correlates with regional case occurrence

The association between COVID-19-confirmed clinical cases and wastewater-N1 or N2 concentrations was tested using a one-sided Fisher’s exact test under the null hypothesis that those two factors were independent for each location (Table 2). An association between cases and a concentration was observed at most monitoring sites (i.e., p<0.05 at RH1, NE, or SO) for samples collected before (−1 week) and the same week (+0 week) using either N1 or N2 as an indicator. As expected, given the mandatory exclusion of confirmed cases from campus following the diagnosis, samples collected the week (+1 week) did not associate.

### 3.5. Wastewater-measured SARS-CoV-2 enabled estimation of COVID-19 cases per capita across UofC campus

COVID-19 cases per capita for the entire UofC monitored catchments comprising NW, NE, and SO during the entire monitoring period (both -A and -B) were estimated according to (Eq. 1) using raw concentrations, and also (Eq. 2) using normalized concentrations. SARS-CoV-2 N2 data was used in this analysis due to its stronger association with clinically confirmed cases (see 3.2, also Table S2). Following this, the modelled aggregate SARS-CoV-2 burden for UofC was compared with the values for the surrounding community (i.e., WWTP catchment) (Fig. 4). For most time points, cases per capita in the community (as measured at WWTP) were significantly higher than for UofC (p<0.001). The results using different methods of potential normalization generally mirrored the raw concentrations (Table 3).

**Figure 4.**
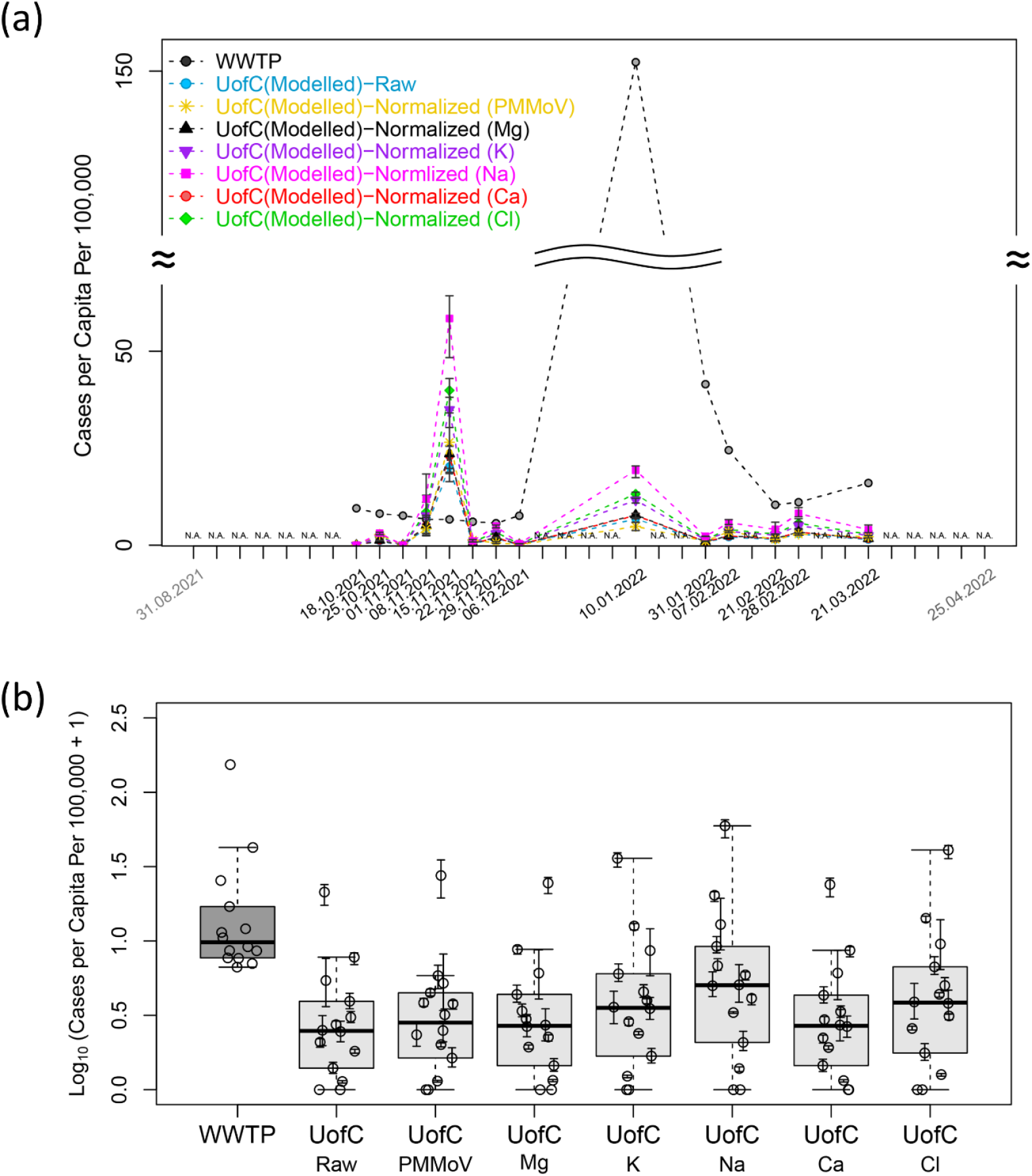
Log_10_-transformed cases per capita estimated for UofC according to Eq.1 (for raw-concentration) or Eq.2 (for normalized-concentrations using PMMoV and chemical agents), and measured for the surrounding community (i.e., WWTP) during the entire monitoring period. Cases per capita was calculated for each data point (i.e., time point), and displayed using trend lines (linearly extrapolated between two data points) over time in (a), and using box plots after aggregated by group in (b). Only paired data points were shown, and statistically compared between each other using Permutation test. Error bars for the modelled (UofC’s) values indicate IQR (Q1-Q3) derived from uncertainty analysis (see 2.7).

Overall, median predicted incident cases per capita (cases per 100,000 scaling factor) for UofC was 5.9-fold lower than for WWTP using raw concentration (p<0.001), and 2.2–5.2-fold lower than for WWTP using normalized concentrations (≤0.01). For instance, the median cases per 100,000 was 8.8 (IQR 6.9-14.8) for the WWTP catchment, and predicted to be 1.5 (IQR 0.5-2.7) for the entire UofC monitoring catchment using raw SARS-CoV-2 RNA concentration. The median values of cases per capita per 100,000 for UofC when assessed using normalized concentrations were 1.8 (IQR 0.7-3.3) for PMMoV, 1.7 (IQR 0.6-3.1) for calcium, 2.9 (IQR 1.0-5.3) for chloride, 1.7 (IQR 0.6-3.1) for magnesium, 2.6 (IQR 0.9-4.7) for potassium, and 4.0 (IQR 1.4-7.6) for sodium.

## 4. Discussion

### 4.1. A nodal-based sampling approach reveals ‘hotspots’ for COVID-19 cases within the campus

This study demonstrated that WBS using spatially resolved node-based sampling approach enables SARS-CoV-2 activity to be located and quantified across a large University campus on a granular scale. Such an approach has previously been proven effective in cities at a neighborhood scale.^11^ One of the challenges for granular scale monitoring is identifying sampling nodes that adequately cover most of the targeted community. This requires a careful analysis of geographic information of involved sewersheds and assessing the connectivity between sampling nodes so that the sub-catchments (for each node) can be collected comprehensively. Other studies have explored SARS-CoV-2 WBS in individual buildings across university campuses. However, those studies either targeted residences and dormitories^16, 18, 37^ or combined residential and non-residential buildings in a very limited fashion.^15, 17, 38^ A comprehensive longitudinal assessment of a campus community or large work facility has not previously been performed. Our approach is unique relative to other studies because our monitoring catchments cover the vast majority of buildings (>83%) on a 530-acres campus through the deployment of a modest number of sample collection devices.

SO and NW portions of the campus consistently showed high levels of SARS-CoV-2 using raw and normalized data, demonstrating hotspots for COVID-19 occurrences. For example, RH1, a dormitory, could be one of the buildings where highest disease incidence occurs and disproportionately contribute to a high level of SARS-CoV-2 signals for NW, at least during Period-B (see Fig. 3). This is consistent with other reports demonstrating high secondary COVID-19 case occurrence in dormitories.^16, 17, 38^ The catchment SO includes three additional residence halls (KA, AU, and RU; see pink sections of Fig. 1), and these may be a reason why SO demonstrated particularly high levels of SARS-CoV-2 signals, especially during Period-B (Fig. 3). Unlike NW and SO, the catchment area for NE does not include any such buildings, rather is predominately comprised of lecture halls and administration offices – this might be one of the reasons why the level of SARS-CoV-2 concentration for NE was low relative to SO or NW. The ability to discern the building(s) with the highest number of cases per capita however, is unknowable in this study. WBS at even more granular scales (i.e., building level) could be followed for those specific building types within catchments of interests, for instance NW and SO in this study, during disease outbreaks, although this would significantly increase the cost of active monitoring by introducing many more nodes.

Comparing the concentration of SARS-CoV-2 RNA in wastewater across different sampling locations, makes it possible to locate the catchment(s) where infected individuals are disproportionally located. However, for such cross-site comparisons, there is a possibility that target analyte abundance could be underestimated in catchment(s) where a high volume of water use relative to individuals is expected (e.g., non-residential buildings). For this reason, we used normalized concentration as a secondary outcome measure when comparing SARS-CoV-2 RNA concentrations across different locations so that we could, in theory, compensate for such underestimation, especially in NE. Other studies have also found that human-specific surrogates did not necessarily improve correlation between confirmed cases and normalized SARS-CoV-2 signals using longitudinal data^39, 40^, similar to our own observations. Another study demonstrated that while normalization using human excretory surrogates did not improve overall correlation of longitudinal data, it still significantly improved the correlation when using “pooled” data for 12 different communities.^41^ This implies that whether normalization improves the correlation between wastewater data and clinical cases depends on site, thus site-specific longitudinal assessments should take precedence.

### 4.2. Campus-wide WBS is positively associated with confirmed COVID-19 case occurrence demonstrating its potential for disease monitoring

A positive association between COVID-19 cases and wastewater signals in the majority of instances (see 3.4 and Table 2) indicates that WBS has the potential for passive disease monitoring at a granular scale. This positive association has previously been reported in other targeted, granular scale monitoring programs. For instance, a positive correlation between wastewater SARS-CoV-2 levels and confirmed cases was observed in hospitals^12, 42^ and university dormitories^38^, and also larger building complexes.^14^ However, adapting WBS as an early warning for COVID-19 cases on a more granular scale may not provide the same lead time relative to clinical diagnoses as was observed early in the pandemic now that testing capacity has markedly increased. Indeed, the early warning scenario (−1 week) did not lead to lower p-values relative to the no time-lag scenario when comparing confirmed COVID-19 cases and wastewater SARS-CoV-2 in this campus monitoring program. A similar observation was reported in another study where node-based sampling strategies were applied for monitoring different neighborhoods at a range of scales (from 853 to 9,094 serving populations) in Illinois, USA.^43^ The authors reported that a correlation between wastewater signals and confirmed cases varied significantly by neighborhood, and an early warning scenario (−1 week) did not necessarily result in a better correlation.^43^

Similar to other studies correlating wastewater measured SARS-CoV-2 with COVID-19 disease occurrence was our reliance on clinically confirmed cases to build models. Individuals with asymptomatic and pauci-symptomatic disease are thusly not captured in this syndromic surveillance-driven manner.^44, 45^ As university campuses generally comprise a younger cohort relative to the general population, the number of asymptomatic infections is expected to have been higher.^46^ Furthermore, as case reporting to University staff was voluntary, it is possible that not all confirmed cases were properly reported. As data was collected by university staff with the primary intent of actionability, cases with missing data (i.e., those with inaccurate dates and details on campus associated movements) were not necessarily followed up on resulting in an incomplete dataset. Finally, the analysis of both wastewater samples and the corresponding campus-confirmed clinical cases were confounded by the use of weekly-aggregate data comparisons. As daily reported cases were discontinuous, and at times relatively low (median=1 and IQR=0-3 for NW; median=0 and IQR=0-1 for NE and SO; med=0 and IQR=0-0 for RH1 and 2), the paired comparison between wastewater signals and reported cases was difficult for campus sites, which is why comparisons were made using weekly-aggregate signals. Daily comparisons would allow for a more accurate analysis of the potential lead time generated through granular WBS, however, such an approach would also create considerable operational and cost challenges.

### 4.3. SARS-CoV-2 activity across University campus was lower than the surrounding community

Our results in Fig. 3 and Fig. 4 demonstrated a much lower viral burden in wastewater across the campus relative to the surrounding community. The relatively low SARS-CoV-2 burden within UofC campus wastewater likely relates to strict COVID-19 mitigating measures mandated within the campus. The ‘COVIDSafe Campus’ run by the university during the pandemic^47^ included mandated proof of vaccination (or documented weekly-negative testing) in order to attend campus, an enforced universal masking mandate and wide availability of hand hygiene product, and a consistent effort for increasing public awareness of COVID-19.

Recent studies have suggested that similar COVID-19 mitigating strategies employed at other university campuses have likewise been effective and that universities were not a large source of disease propagation. For instance, a SARS-CoV-2 phylogenetic study performed at the University of Michigan, USA, revealed that the descendants of SARS-CoV-2 from student cases were rarely found in the community during the next wave.^48^ In another study performed at the University of Cambridge, UK, the authors revealed that the majority of SARS-CoV-2 genomes from students originated from a single genetic cluster – the cases occurred after a single event (e.g., social gathering outside the campus), suggesting a limited introduction of the virus into the community.^49^ Likewise, we did not observe new VOC occur earlier within the campus than the general community. Collectively these studies suggest that the intensive efforts to reduce forward transmission of COVID-19 adopted in higher education settings could be applied to other contexts to mitigate further disease spread in those environments.

### 4.4. Other notable limitations

There are several other noteworthy limitations of this study. For instance, toileting patterns do vary considerably across space and time. In particular, there have been reports documenting that many individuals prefer to defecate at home^50^, and these active cases would therefore be underrepresented in work-based studies. Accordingly, work (or school)-based studies such as those monitoring campuses may underestimate the true burden of infected populations within. We attempted to mitigate for this factor by assessing SARS-CoV-2 RNA concentration both raw, and normalized against fecal and population surrogates, where we observed the same general trends.

Furthermore, the monitoring in this study was performed when not all students and employees had fully returned to in-person learning/work; a small number continued to telecommute from home and, therefore, may not fully represent the entire university community. Thus, care has to be taken when interpreting our results – the results in this study do not indicate for instance that university members tend to have lower disease infection rates then populations outside the campus, but rather suggest that university campus is not the place where high cases per capita exist, or diseases were contained relatively well ‘within the campus’. Finally, wastewater-based monitoring at a granular scale may not fully represent actual case burden within the catchment because individual confounding differences may have a larger effect relative to community monitoring. Viral shedding may vary by individual^51^ and the chances of capturing shedding events when autosamplers were being operated are highly stochastic, etc. We attempted to address this issue by employing 24-hours composite sampling over a “grab” sampling strategy, and by achieving a reasonably high sample size (i.e., up to 35 points for weekly aggregated signals from 89 individual data points) followed by various statistics (e.g., non-parametric tests such as Kruskal-Wallis, Wilcoxon tests). In this way, our wastewater results could provide an “approximate” to the case per capita existing in each monitoring catchment.

To our knowledge, this is the first study to comprehensively assess SARS-CoV-2 (and VOC) burden across a large university campus using a spatially resolved, nodal based strategy. We have confirmed the potential of this platform technology to perform population health monitoring through wastewater analysis. This study has established wastewater-based surveillance is positively associated with clinical cases at a granular scale, suggesting it can be used synergistically with contact tracing in order to identify ‘hotspots’ for COVID-19 occurrence across campus (i.e., building). This study also confirmed the markedly lower rates of SARS-CoV-2 across campus, lending support to the importance of restrictive measures in mitigating COVID-19’s potential for spread across worksites.

## Data Availability

Key data produced in the present work are contained in the manuscript. The other data (not-shown) produced in the present study are available upon reasonable request to the authors.

## Acknowledgements

This study was supported by Alberta Health and the Canadian Institutes of Health Research. Dr. Jangwoo Lee received salary support from a University of Calgary Eyes High Fellowship. The authors are grateful for the assistance of the staff of City of Calgary Water Services and C.E.C. analytics for training assistance in sewershed access and sample collection.

