## Supplementary Information_Methods, Figures, and Tables for "Spatially-resolved wastewater-based surveillance enables COVID-19 case localization across a university campus, and confirms lower SARS-CoV-2 RNA burden relative to the surrounding community"

### 1. Supplementary Methods

#### 1.1 Field sampling

For the two sample locations capturing the University residences (RH1 and RH2), C.E.C Analytics V1 autosamplers (C.E.C Analytics, Canada) were installed and programmed to collect wastewater 2 times a week (Monday, and Wednesdays) from August 31^st^ until November 17^th^, and three times a week (Monday, Wednesday, and Thursday) from November 18^th^ until April 24^th^. A total volume of 2L was collected for each wastewater sample using 4 x 500 mL bottles connected in series, the excess liquid fraction drained from the series back into the sewerline. At the lab, the wastewater from all four bottles was mixed thoroughly before an aliquot was taken for the purpose of analysis.

The three outdoor locations (NE, SO and NW) were initially installed in municipal sewershed in late September with a Teledyne ISCO 6712 autosampler (Teledyne ISCO, USA) and programmed to collect 100ml of wastewater every 15 minutes for 24 hours, initially twice weekly, Mondays, and Wednesdays and then 3 times weekly from November 18^th^ forward. The ISCO collected a total volume of 9.6L which was mixed thoroughly at the lab before an aliquot was taken for analysis. The three outdoor locations were shifted over to a suspended in-sewer C.E.C Analytics autosamplers due to winter conditions to protect against freezing. The C.E.C Analytics autosamplers were programmed and samples handled in the same manner as the residence samples.

To establish equipoise between sample collection devices, we performed a three-week cross-over comparison between the ISCO and the C.E.C. autosamplers where duplicate samples were collected from each device. For this comparison, a total of 38 samples (from November 15 to December 6) were obtained from two sites, which were not included in this project, but obtained for other monitoring projects on-going in parallel. The data (two SARS-CoV-2 markers (N1 and N2), a fecal marker (PMMoV, Pepper Mild Mottle Virus), and a spike control (BCoV, Bovine Coronavirus)) obtained using those two autosamplers significantly correlated with each other (r = 0.85, 0.78, 0.92, and 0.88 for N1, N2, PMMoV, and BCoV respectively, all p<0.001), and did not significantly differ using Wilcoxon test (p>0.05), which confirmed that data derived from samples collected with both devices are comparable.

#### 1.2. 4S- Sample processing and RNA extraction details

After being passed through a Zymo-spin column, two consecutive washes were performed. First, an excess wash buffer was removed via centrifugation at 10,000 G for two minutes. Next, elution was performed using 100 µL of molecular grade water heated to 50°C, added to the silica column and incubated for two minutes at 50°C. A final centrifugation at 10,000 G for five minutes was performed for nucleic acid recovery. A more detailed description can be found in Whitney et all., 2021^23^. A total volume of 40mL was processed for each sample. Samples were either entirely wastewater or were diluted with Ultrapure molecular grade water. Samples were diluted according to visual inspection based on solid levels and color and transparency of the liquid fraction, with downstream analyses accounting for the dilution (a dilution of 1:2 was most commonly used).

#### 1.3. Fecal-normalized concentration of SARS-CoV-2 as an indicator of infections per capita

The concentration of SARS-CoV-2 RNA in wastewater in a defined catchment relates to the number of actively infected individuals. For example, the total SARS-CoV-2 RNA in the wastewater equals the summation of SARS-CoV-2 RNA discharged from people with COVID-19 existing in the catchment area at a specific time interval.

$C\cdot V=\sum_{i=1}^{n} L_{p_{i}} (Eq. S1)$

Where $C$ indicates the concentration of SARS-CoV-2 RNA in the wastewater; $V$ denotes the volume of wastewater; $L_{p}$ indicates SARS-CoV-2 loading per person; $n$ indicates the total number of COVID-19 occurrences in a catchment area

Under the following two assumptions (Assumptions 1 and 2), the equation above (Eq. 1) could be reduced to (Eq. 4).

*Assumption 1: Every individual contributes equally to wastewater SARS-CoV-2 signal (i.e.,* $v$ *is a constant),* which could be summarized as the following equation.

$$V=N\cdot v (Eq. S2)$$

where $N$ is the number of individuals in the catchment area; $v$ denotes the volume of individual water usage, and is a constant.

Also,

$$\sum_{i=1}^{n} L_{p_{i}}=v\cdot\sum_{i=1}^{n} c_{i} (Eq. S3)$$

Where $c_{i}$ indicates the concentration of SARS-CoV-2 RNA in the feces discharged from each person with COVID-19

*Assumption 2: SARS-CoV-2 shedding does not significantly vary between individuals with COVID-19*

On Following the assumptions that i) most shedding (hereby defined as concentration) occurs during the first few days of disease onset ^12, 22^ (i.e. ~2 days before or after the date of onset of symptoms), ii) this does not significantly vary among individuals, or at least follows certain probability distributions at a population level, thus sum of viral shedding could be approximated to

$$\sum_{i=1}^{n} c_{i} \approx\bar{c}\cdot n$$

where $\bar{c}$ indicates a representative value (e.g., average, median, etc) for wastewater SARS-CoV-2 RNA concentration for a defined population

The variables $V$and $\sum_{i=1}^{n} L_{p_{i}}$ from (Eq. 1) could be substituted to (Eq. 2) and (Eq. 3) as follows, and the final reduced form is suggested in (Eq. 4).

$$C\cdot N\cdot v=\sum_{i=1}^{n} L_{p_{i}}=v\cdot\sum_{i=1}^{n} c_{i}=\bar{c}\cdot n\cdot v$$

$\therefore C=\bar{c}\cdot\frac{n}{N}$	or $C \propto\frac{n}{N}$	(Eq.S4)

The (Eq. 4) implies that the concentration of SARS-CoV-2 RNA in the wastewater ($C$) is proportional to the number of COVID-19 infected individuals with COVID-19 per capita ($\frac{n}{N}$) in a defined catchment area.

However, $\bar{c}$ may be a population specific term, so this may be variable depending on population (or catchment). For instance, $\bar{c}$ may be lower for a catchment where individual water usage is higher due to dilution (e.g., more water consumption due to research or dining activities, as opposed to defecation). Therefore, what could be more constant is “$\bar{c}$ relative to the concentration of human excreta ($\bar{c_{e}}$) in water discharged from individual” rather than $\bar{c}$.

$$\frac{C}{\bar{c_{e}}}=\bar{\frac{c}{\bar{c_{e}}}}\cdot\frac{n}{N}$$

$\bar{c_{e}}$ could be approximated to the human excreta concentration in the (pooled) wastewater ($C_{e}$) according to the following rationale.

Assuming that $\bar{m}\cdot N\approx\sum_{1}^{N} m_{i}$

$$\bar{c_{e}}=\frac{\bar{m}}{v}=\frac{\bar{m}\cdot N}{v\cdot N}=\frac{\sum_{1}^{N} m_{i}}{V}=C_{e}$$

Where, $m$ indicates a mass (or copy) for virus shedding from individual; $\bar{m}$ denotes a representative value for $m$.

Therefore,

$\frac{C}{C_{e}}=a\cdot\frac{n}{N}$	(Eq.5)

Where, $a= \frac{c}{\bar{c_{e}}}$, and a constant across populations

#### 1.4. Modelling expected COVID-19 cases per capita across UofC Campus

Given that our monitoring catchments cover the vast majority of the UofC main campus, the total loading of wastewater-borne virus for the entire campus ($L_{UofC}$) could be approximated to the summation of viral load ($L_{SO}, L_{NW}, or L_{NE}$) from all three campus catchments.

$$L_{UofC}\approx L_{SO}+ L_{NW}+ L_{NE}$$

$$C_{UofC}\cdot V_{UofC} \approx C_{SO}\cdot V_{SO}+ C_{NW}\cdot V_{NW}+ C_{NE}\cdot V_{NE}$$

Where, $C$ indicates viral concentration; $V$ denotes the volume of wastewater

The following relationship is met under the assumption that the flow quantity of wastewater is proportional to the surface area of the catchment ($A_{SO or NW or NE}$). The catchment surface area was defined as a summation of gross area (m^2^) for all the buildings belonging to the catchment. On the other hand, NW and NE have six buildings which overlap – the common shared sub-catchment ($A_{MIX}$; orange colored in Fig. 1) should be subtracted from either NW or NE (subtracted from NE in this case).

$$V_{SO} : V_{NW} : V_{NE-MIX}=A_{SO} :A_{NW} :A_{NE-MIX}$$

Where, $A_{SO}$ = 83,884 m^2^, $A_{NW}$ = 440,610 m^2^, and $A_{NE-MIX}$ = 105,504 m^2^

Assuming that $V_{tot}$ is approximated to the summation of wastewater flow quantities for all three catchments ($V_{UofC}\approx V_{SO}+V_{NW}+V_{NE-MIX}$), the following equation could be derived.

We assumed $C_{NE-MIX}$could be approximated to $C_{NE}$ ($C_{NE-MIX}\approx C_{NE}$) for the following rationales: i) both the shared area ($A_{MIX}$) and the rest of NE consisted of similar types of buildings (non-residential and non-recreational buildings) – wastewater characteristics, and also the probability of COVID-19 occurrence may be similar, and ii) the common area consisted of a significant portion (51%) of NE.

Therefore,

$$C_{UofC}\cdot\left( A_{SO}+A_{NW}+A_{NE-MIX} \right)\cdot a=(A_{SO}\cdot{nC}_{SO}+A_{NW}\cdot C_{NW}+A_{NE-MIX}\cdot C_{NE})\cdot a$$

$\therefore C_{UofC}=\frac{A_{SO}\cdot C_{SO}+A_{NW}\cdot C_{NW}+A_{NE-MIX}\cdot C_{NE}}{A_{SO}+A_{NW}+A_{NE-MIX}}$	(Eq.S6)

Where, $a$ indicates a proportional constant.

#### 1.5. Correlating clinically confirmed cases with regional SARS-CoV-2 wastewater measured RNA

We hypothesized that the COVID-19 clinically-confirmed cases (Var-1) within a sewershed catchment may be positively related to the wastewater SARS-CoV-2 signal measured (Var-2), and also serve as an early warning indicator. We therefore performed a Fischer’s exact test between those two variables (i.e., Var-1 and -2) under each of the following assumptions:

**Assumption-1** : An early warning relationship between Var-1 and -2 (i.e. week-1)

**Assumption-2** : No time lag relation between Var-1 and -2

**Assumption-3** : A time lagged relationship between Var-1 and -2 (i.e. week+1)

According to the abovementioned assumptions, Var-1 and -2 were paired differently, for instance Var-2 was paired with a week early data (-1 week) for Var-1 (**Assmption-1**), with Var-1 for each corresponding time point (**Assumption-2**), and with a time lagged data (+1 week) for Var-1 (**Assmption-3**). This was graphically abstracted as follows (Fig. S1).

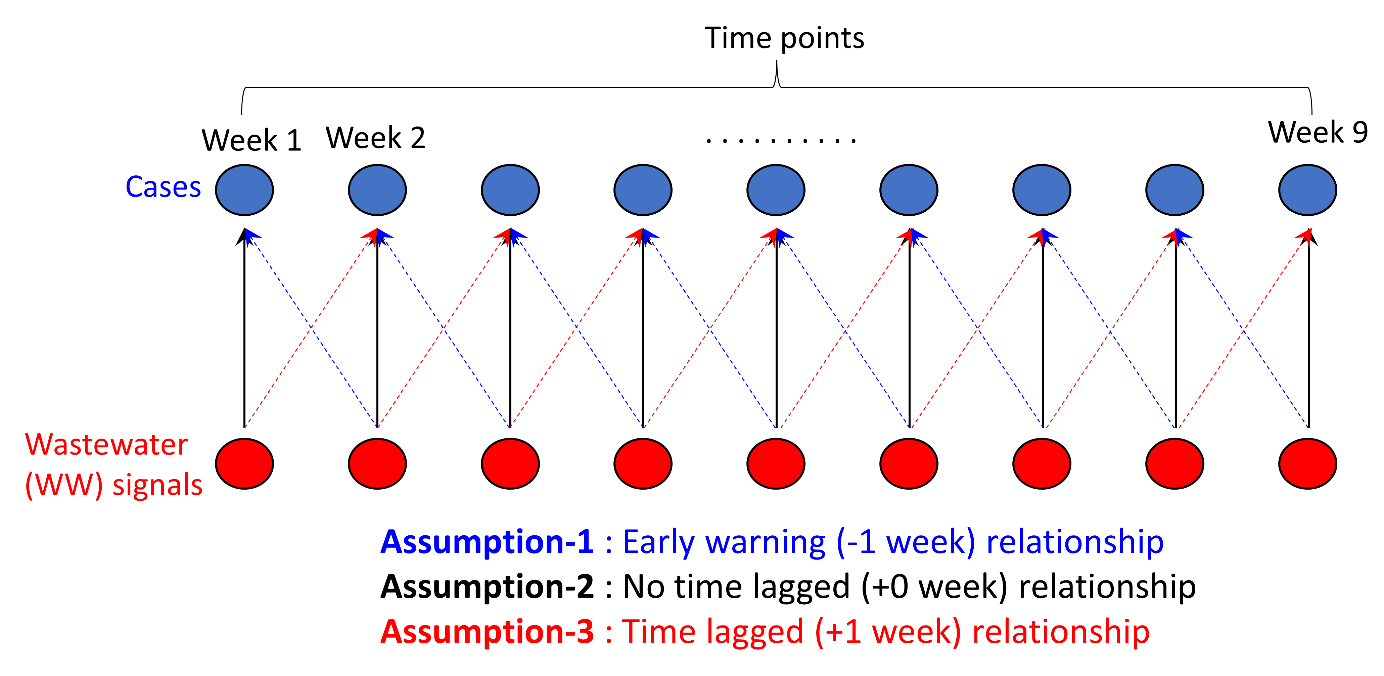

**Figure S1**. Data pairing between COVID-19 clinically confirmed cases in a particular catchment area and wastewater measured SARS-CoV-2 RNA for each assumption.

We then used a contingency table to test for each assumption (-1 to -3) per monitoring location. For instance, i) levels of each measured variable was defined as high (> Q2, 2^nd^ quantile range) and low (≤ Q2), ii) the frequency distribution of the variables was summarised in a 2 x 2 table as follows (see Fig. S2), iii) the steps i) – ii) were repeated for each assumption (-1 to -3; see Fig. S1) for each monitoring location (i.e., RH1, RH2, SO, NW, or NE). This resulted in a total of three contingency tables per location.

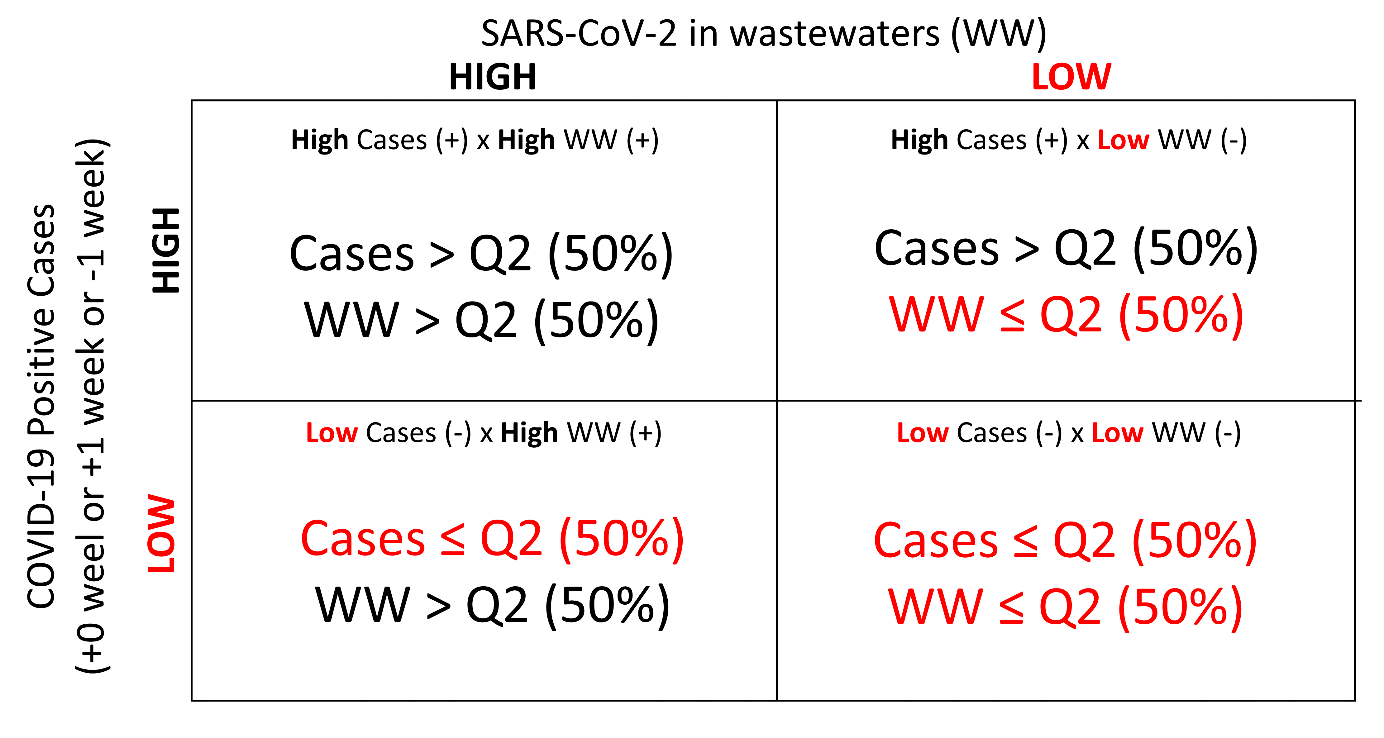

**Figure S2**. A schematic example of contingency table produced for each assumption.

Finally, Fischer’s exact test was performed for each table, and all the results wer summarized in Table 1. One-side test was employed under the assumption that Var-1 and -2 were ‘positively’ related. The lower the p-value is, the more dependent the two variables are.

### 2. Supplementary Results

#### 2.1 A brief summary of protocol for quality control workflows (for RT-qPCR data)

All exogenous spiked BCoV values were higher than the minimum acceptable predetermined values (> 62,500 copies/mL) (Acosta et al., 2021; Acosta et al., 2022) and therefore no samples were excluded on this basis. For instance median BCoV (IQR) was 3,451,102 (1,519,389-8,362,931) copies/mL for RH1, 1,115,715 (409,182-3,714,629) copies/mL for RH2, 1,230,803 (723,711-2,289,143) copies/mL for NE, 2,861,288 (1,198,225-5,577,698) copies/mL for SO, 1,535,941 (767,179- 2,812,140) copies/mL for NW, and 1,274,963 (747,273-2,186,352) copies/mL for WWTP.

Outliers were ientified referring to PMMoV signals, for instance the samples with too high (i.e., > 3^rd^ quantile (Q3) + 1.5 × Interquartile Range (IQR)) or too low (i.e., < 1^st^ quantile (Q1) – 1.5 × IQR) signals for PMMoV were considered as outliers (a total of 2 out of 58 samples) containing too much or low amounts of human feces (see Fig. S3) – those values were removed from further downstream analysis.

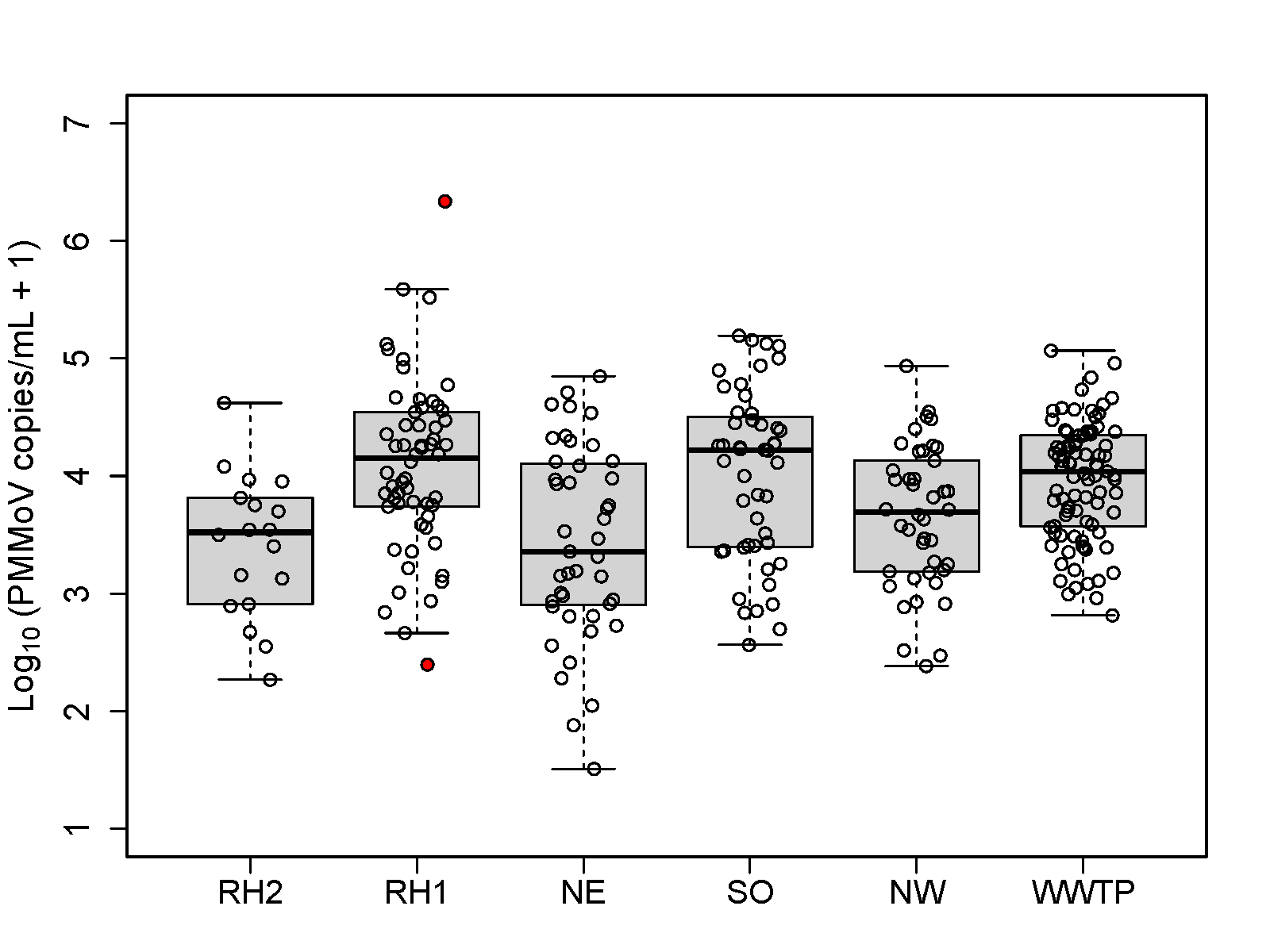

**Figure S3.** Log_10_-transformed abundances (copies/mL) of Pepper Mild Mottle Virus (PMMoV) for each monitoring locations during the entire monitoring period, -A and -B. RH1 or 2 indicates Residence Hall 1 or 2, and NE, NW, and SO denotes Northeastern, Northwestern, and Southern campus districts. WWTP indicates wastewater treatment plant. The outliers excluded from analysis (> Q3 + 1.5 × IQR or < Q1 - 1.5 × IQR) are highlighted in red.

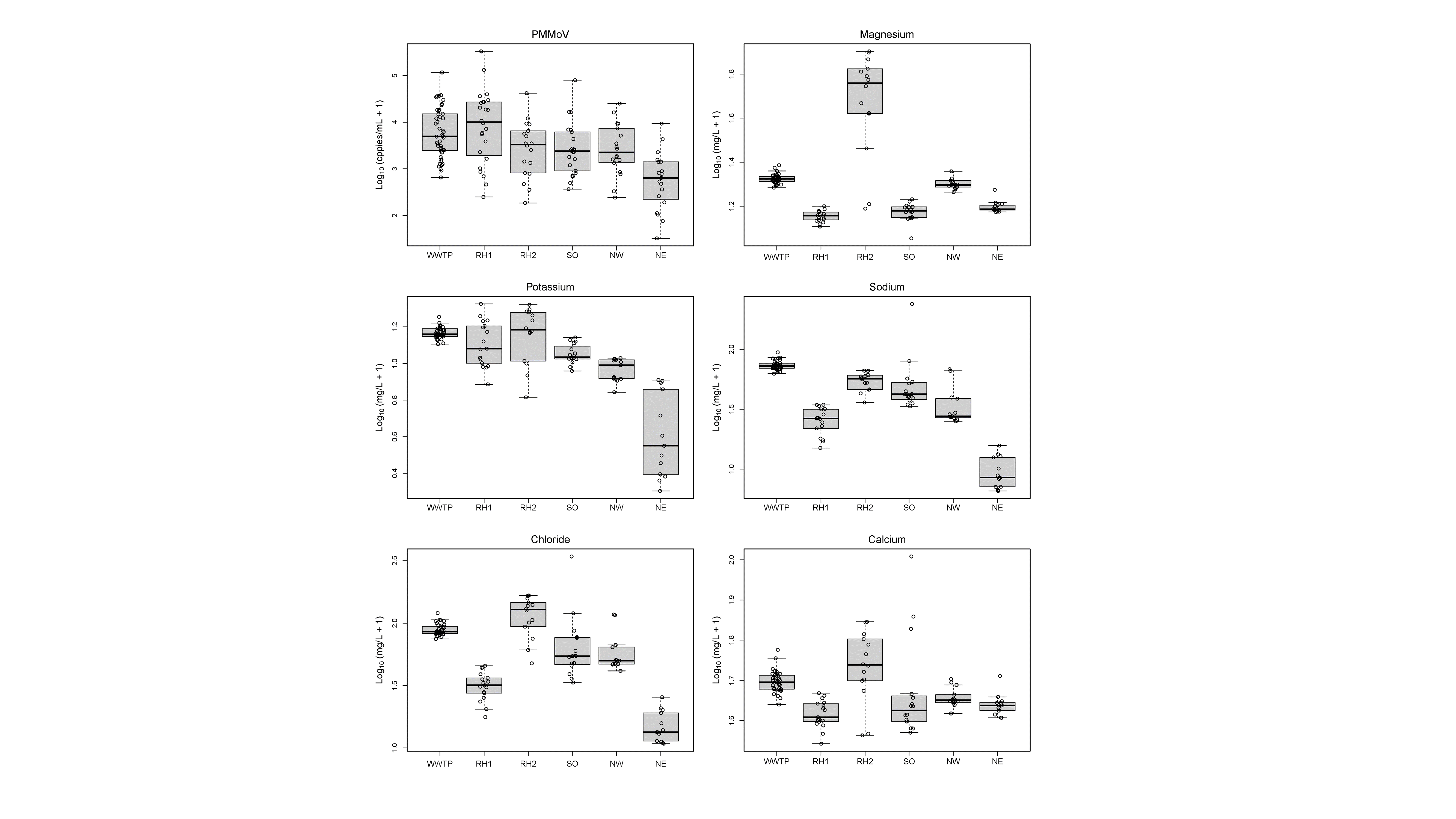

#### Figure S4. Concentrations of potential human activity normalization markers (PMMoV, potassium, sodium, chloride, calcium, and magnesium) by monitoring location (date range : August 2021 – January 2022; n = 13 – 36 depending on location). RH1 or 2 indicates Residence Hall 1 or 2, and NE, NW, and SO denotes Northeastern, Northwestern, and Southern campus districts. WWTP indicates wastewater treatment plant.

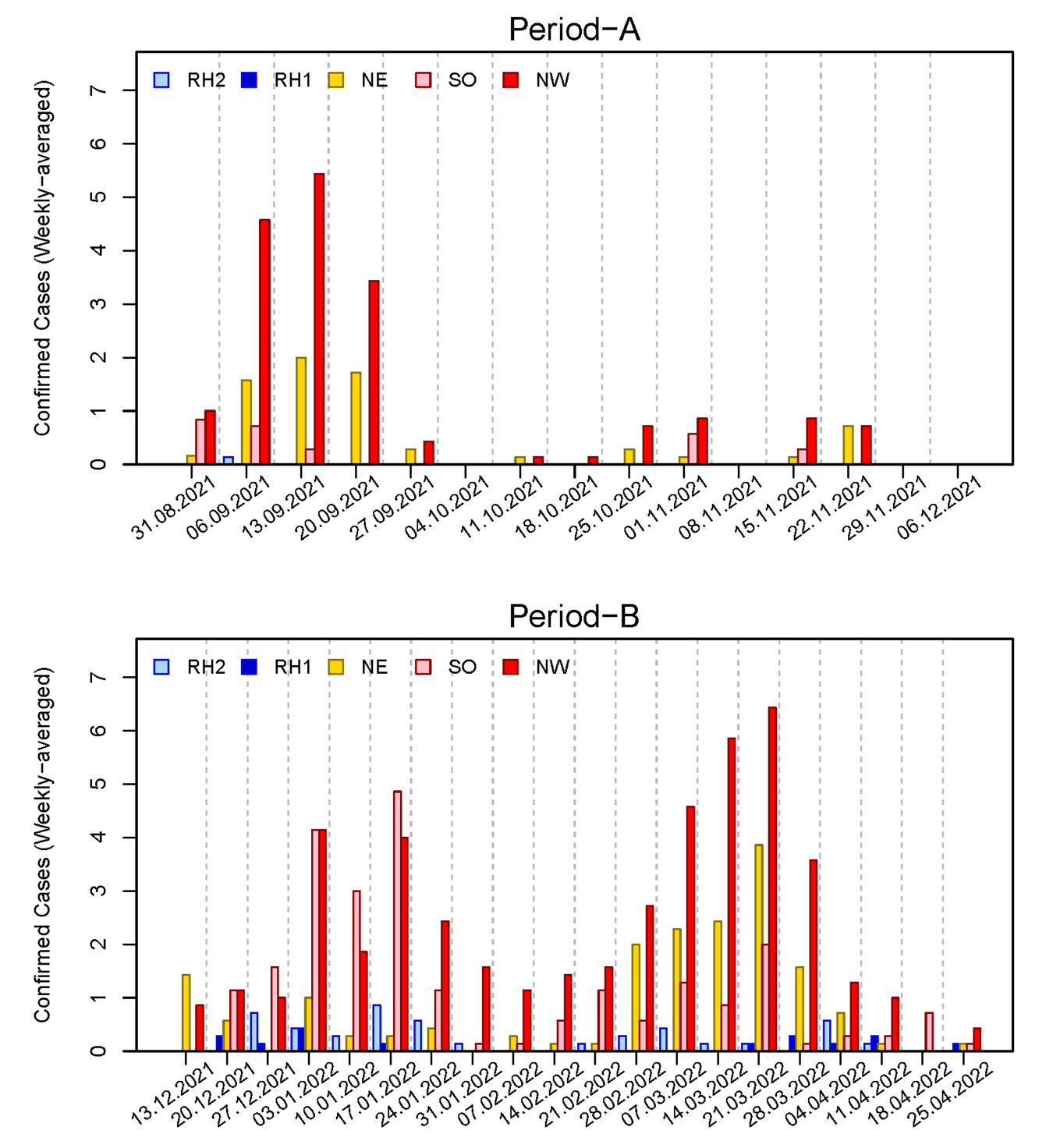

#### Figure S5. Confirmed cases (weekly averaged) for each monitoring catchment during Period-A (upper) and -B (lower). RH1 or 2 indicates Residence Hall 1 or 2, and NE, NW, and SO denotes Northeastern, Northwestern, and Southern campus districts.

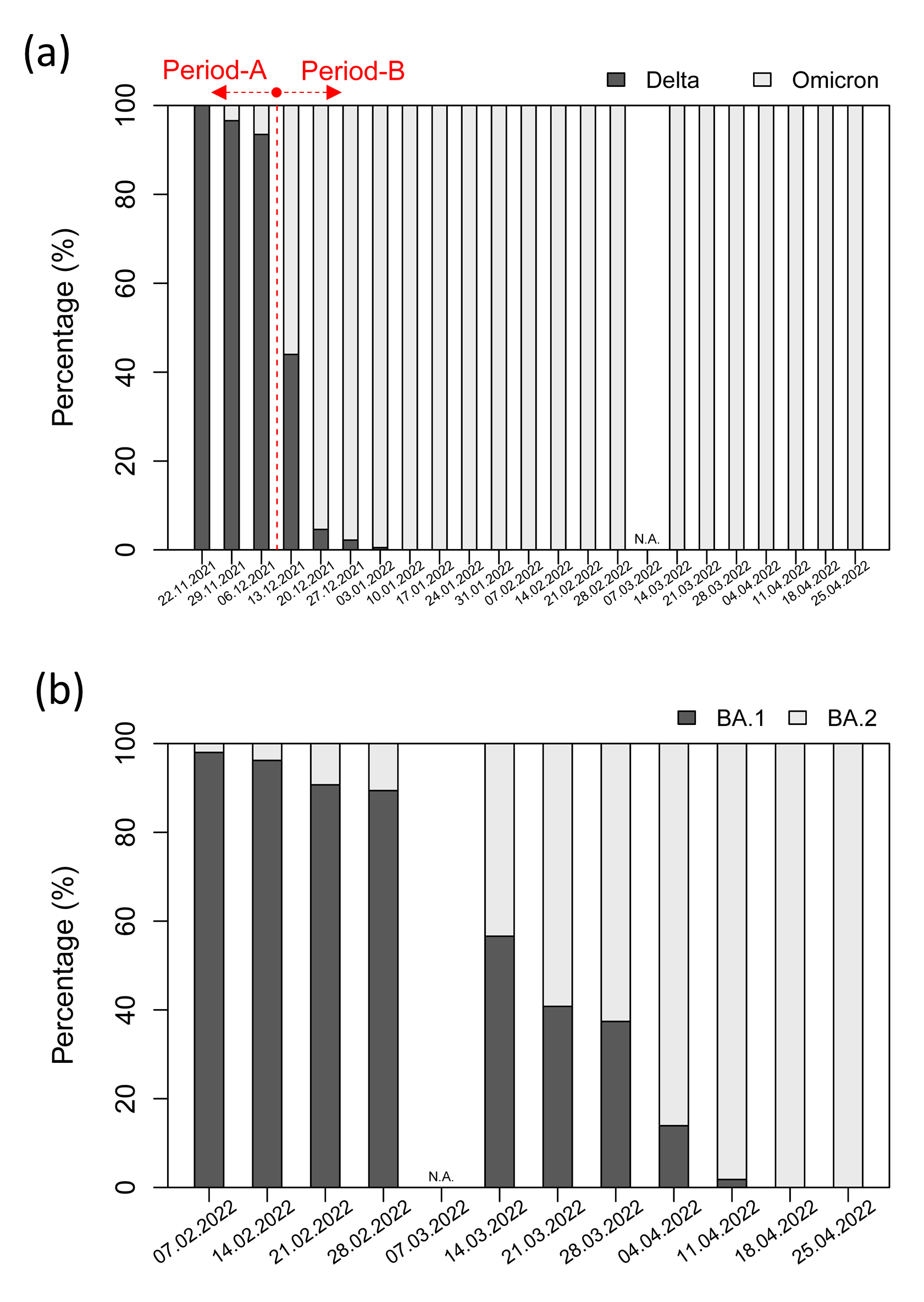

#### Figure S6. The relative proportion (in percentage) of (a) Delta to Omicron BA.1 lineage, and (b) Omicron BA.1 to BA.2 lineage of SARS-CoV-2 identified in this study for wastewater treatment plant (receiving wastewaters from UofC campus, also from the surrounding community) during a subset of monitoring period (22.11.2021 – 25.04.2022 for (a); 07.02.2022 – 25.04.2022 for (b) – belong to Period-B).

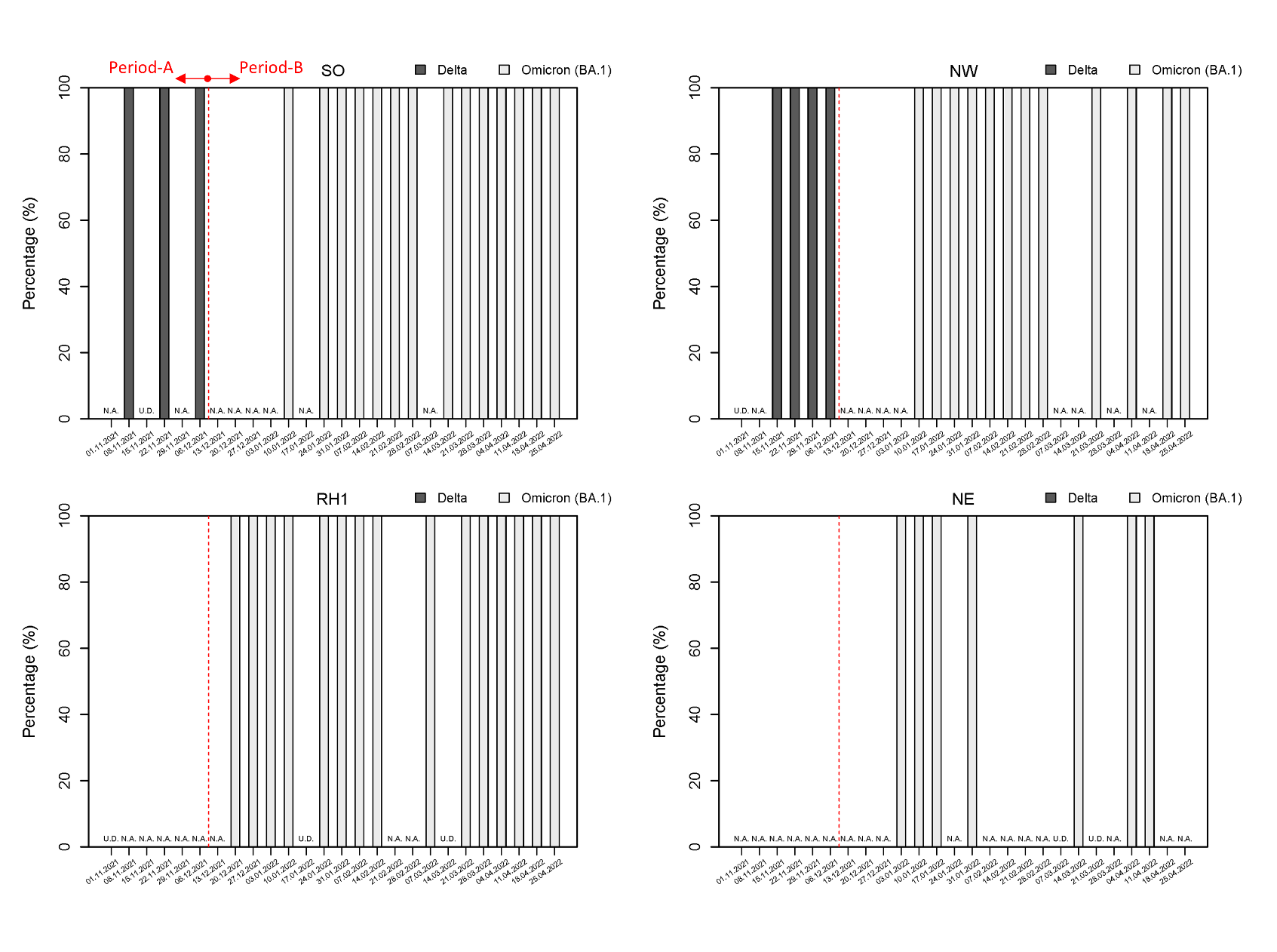

#### Figure S7. The relative proportion (in percentage) of Delta to Omicron BA.1 lineage during a subset of monitoring period (01.11.2021 – 28.04.2022) for each UofC campus monitoring location. RH1 indicates Residence Hall 1, and NE, NW, and SO denotes Northeastern, Northwestern, and Southern campus districts.

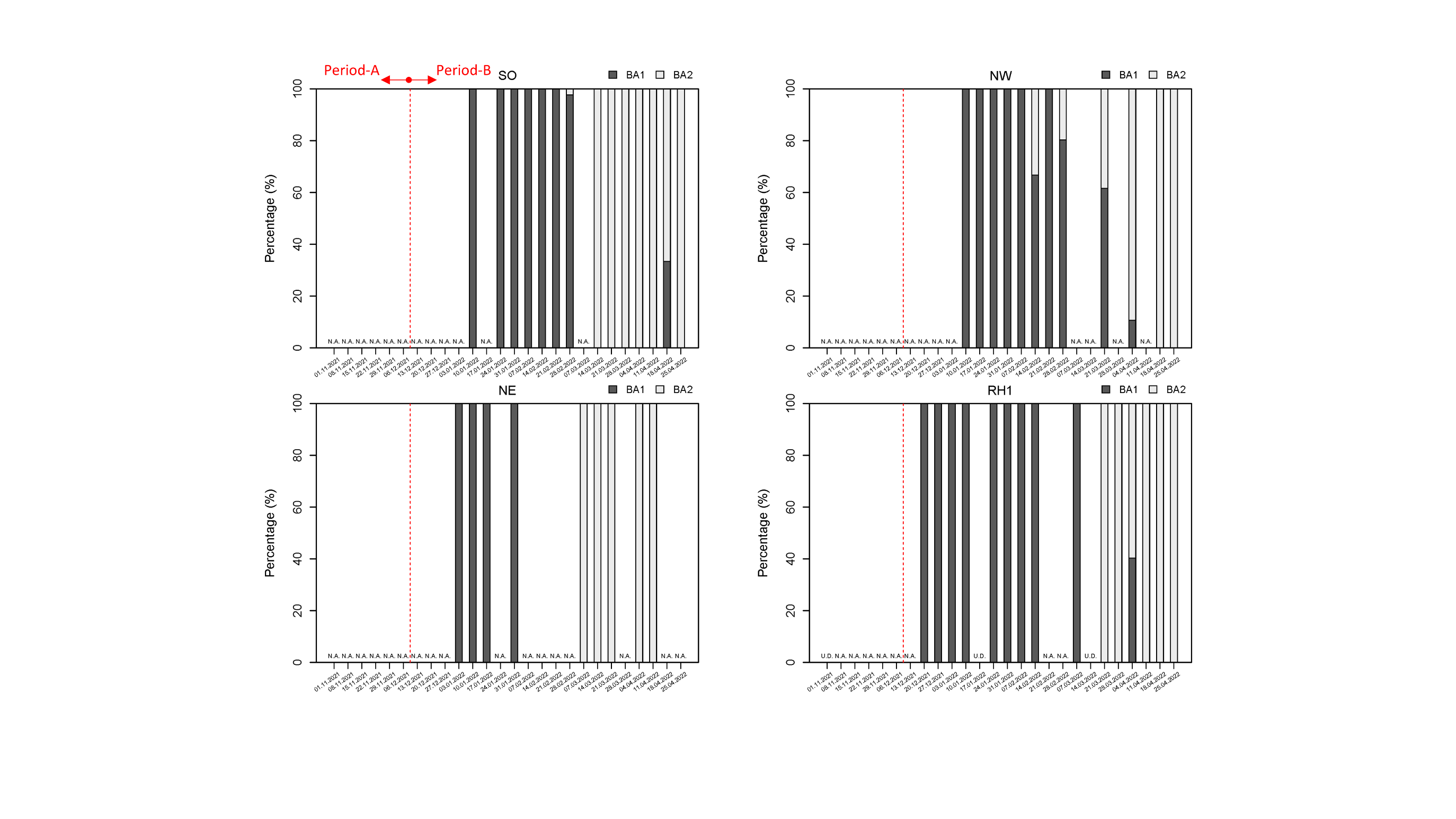

#### Figure S8. The relative proportion (in percentage) of Omicron BA.1 to BA.2 lineage of SARS-CoV-2 during a subset of monitoring period (01.11.2021 – 28.04.2022) for each UofC campus monitoring location. RH1 indicates Residence Hall 1, and NE, NW, and SO denotes Northeastern, Northwestern, and Southern campus districts.

#### Table S1. Key parameters for qPCR standard curves in this study. Median and Interquartile range (IQR) values were suggested below except for *N200 assay where lower – upper values between two plate runs were suggested.

| Marker | Efficiency  (Median (IQR)) | R^2^  (Median (IQR)) | Y-Intercept  (Median (IQR)) | Slope  (Median (IQR)) | Number of Plates |
| --- | --- | --- | --- | --- | --- |
| N1 | 107.6  (100.8-113.7) | 0.98  (0.97-0.99) | 39.6  (39.0-40.1) | -3.15  (-3.30 – -3.03) | 87 |
| N2 | 93.3  (87.6-101.7) | 0.97  (0.96-0.98) | 42.8  (42.2-43.7) | -3.49  (-3.66 – -3.28) | 87 |
| BCoV | 94.4  (87.9-106.1) | 0.99  (0.98-1.00) | 41.1  (38.8-42.7) | -3.46  (-3.65 – -3.18) | 86 |
| PMMoV | 98.2  (90.9-110.0) | 0.99  (0.97-1.00) | 43.6  (41.3-45.3) | -3.36  (-3.56 – -3.1) | 87 |
| *Omicron BA.1 (N200 Assay) | 95.6-96.7  (-) | 0.99-1.00  (-) | 39.0-41.1  (-) | -3.43 – -3.40  (-) | 2 |
| *Delta (N200 Assay) | 95.5-97.8  (-) | 0.98-0.99  (-) | 40.0-40.3  (-) | -3.43 – -3.38  (-) | 2 |
| Omicron BA.1 (60/70del) | 94.5  (93.2-97.5) | 1.00  (0.99-1.00) | 41.6  (41.3-43.1) | -3.46  (-3.50 – -3.38) | 7 |
| Omicron BA.2 (60/70del) | 94.0  (91.2-95.0) | 1.00  (0.99-1.00) | 40.0  (39.2-40.2) | -3.47  (-3.55 – -3.45) | 7 |

#### Table S2. Results for Spearman correlation between clinically confirmed cases and SARS-CoV-2 signals (N1/N2) with or without normalization in wastewaters from a community wastewater treatment plant (date range : August 31 2021 – January 04 2022; a total of 36 data points).

| Normalization Agents | N1 | | N2 | |
| --- | --- | --- | --- | --- |
|  | r | p-value | r | p-value |
| PMMoV | 0.262 | 0.123 | 0.294 | 0.082 |
| Sodium | 0.796 | < 0.001 | 0.801 | < 0.001 |
| Chloride | 0.790 | < 0.001 | 0.799 | < 0.001 |
| Potassium | 0.798 | < 0.001 | 0.825 | < 0.001 |
| Calcium | 0.798 | < 0.001 | 0.821 | < 0.001 |
| Magnesium | 0.802 | < 0.001 | 0.822 | < 0.001 |
| Raw (without normalization) | 0.804 | < 0.001 | 0.823 | < 0.001 |

#### Table S3. The p-values for comparing SARS-CoV-2 RNA normalized concentrations between different monitoring locations during Period-A using Wilcoxon rank-sum test. P-adjusted for pairwise comparison using Benjamini & Hochberg method. Those pairs that statistically differed with (p < 0.05) are highlighted in red.

| **Normalized by** | **Indicator** | **Location** | **RH2** | **RH1** | **NE** | **SO** | **NW** |
| --- | --- | --- | --- | --- | --- | --- | --- |
| Magnesium | N1 | RH1 | 0.690 | - | - | - | - |
|  |  | NE | 0.689 | 0.650 | - | - | - |
|  |  | SO | 0.011 | 0.012 | 0.018 | - | - |
|  |  | NW | 0.007 | 0.009 | 0.008 | 0.916 | - |
|  |  | WWTP | 0.000 | 0.000 | 0.000 | 0.008 | 0.002 |
|  | N2 | RH1 | 0.475 | - | - | - | - |
|  |  | NE | 0.944 | 0.428 | - | - | - |
|  |  | SO | 0.039 | 0.020 | 0.068 | - | - |
|  |  | NW | 0.035 | 0.015 | 0.040 | 0.794 | - |
|  |  | WWTP | 0.000 | 0.000 | 0.000 | 0.002 | 0.002 |
| Potassium | N1 | RH1 | 0.597 | - | - | - | - |
|  |  | NE | 0.597 | 0.373 | - | - | - |
|  |  | SO | 0.024 | 0.012 | 0.085 | - | - |
|  |  | NW | 0.012 | 0.007 | 0.017 | 0.597 | - |
|  |  | WWTP | 0.000 | 0.000 | 0.000 | 0.009 | 0.007 |
|  | N2 | RH1 | 0.475 | - | - | - | - |
|  |  | NE | 0.944 | 0.428 | - | - | - |
|  |  | SO | 0.052 | 0.020 | 0.068 | - | - |
|  |  | NW | 0.035 | 0.015 | 0.044 | 0.679 | - |
|  |  | WWTP | 0.000 | 0.000 | 0.000 | 0.002 | 0.003 |
| Sodium | N1 | RH1 | 0.644 | - | - | - | - |
|  |  | NE | 0.539 | 0.434 | - | - | - |
|  |  | SO | 0.032 | 0.029 | 0.221 | - | - |
|  |  | NW | 0.013 | 0.013 | 0.085 | 0.539 | - |
|  |  | WWTP | 0.000 | 0.000 | 0.000 | 0.013 | 0.013 |
|  | N2 | RH1 | 0.475 | - | - | - | - |
|  |  | NE | 0.944 | 0.428 | - | - | - |
|  |  | SO | 0.052 | 0.020 | 0.083 | - | - |
|  |  | NW | 0.035 | 0.015 | 0.052 | 0.624 | - |
|  |  | WWTP | 0.000 | 0.000 | 0.000 | 0.002 | 0.006 |
| Chloride | N1 | RH1 | 0.644 | - | - | - | - |
|  |  | NE | 0.528 | 0.528 | - | - | - |
|  |  | SO | 0.027 | 0.027 | 0.221 | - | - |
|  |  | NW | 0.008 | 0.017 | 0.085 | 0.640 | - |
|  |  | WWTP | 0.000 | 0.000 | 0.000 | 0.008 | 0.005 |
|  | N2 | RH1 | 0.475 | - | - | - | - |
|  |  | NE | 0.944 | 0.428 | - | - | - |
|  |  | SO | 0.052 | 0.020 | 0.083 | - | - |
|  |  | NW | 0.035 | 0.015 | 0.052 | 0.679 | - |
|  |  | WWTP | 0.000 | 0.000 | 0.000 | 0.002 | 0.003 |
| Calcium | N1 | RH1 | 0.675 | - | - | - | - |
|  |  | NE | 0.839 | 0.650 | - | - | - |
|  |  | SO | 0.026 | 0.014 | 0.020 | - | - |
|  |  | NW | 0.014 | 0.008 | 0.006 | 0.748 | - |
|  |  | WWTP | 0.000 | 0.000 | 0.000 | 0.006 | 0.002 |
|  | N2 | RH1 | 0.475 | - | - | - | - |
|  |  | NE | 0.944 | 0.428 | - | - | - |
|  |  | SO | 0.052 | 0.020 | 0.068 | - | - |
|  |  | NW | 0.038 | 0.015 | 0.038 | 0.736 | - |
|  |  | WWTP | 0.000 | 0.000 | 0.000 | 0.002 | 0.003 |
| PMMoV | N1 | RH1 | 0.655 | - | - | - | - |
|  |  | NE | 0.655 | 0.500 | - | - | - |
|  |  | SO | 0.029 | 0.021 | 0.085 | - | - |
|  |  | NW | 0.008 | 0.017 | 0.021 | 0.655 | - |
|  |  | WWTP | 0.000 | 0.000 | 0.000 | 0.008 | 0.008 |
|  | N2 | RH1 | 0.475 | - | - | - | - |
|  |  | NE | 0.944 | 0.428 | - | - | - |
|  |  | SO | 0.048 | 0.020 | 0.083 | - | - |
|  |  | NW | 0.028 | 0.015 | 0.064 | 0.736 | - |
|  |  | WWTP | 0.000 | 0.000 | 0.000 | 0.005 | 0.015 |

#### Table S4. The p-values for comparing SARS-CoV-2 RNA normalized concentrations between different monitoring locations during Period-B using Wilcoxon rank-sum test. P-adjusted for pairwise comparison using Benjamini & Hochberg method. Those pairs that statistically differed with (p < 0.05) are highlighted in red.

| **Normalized by** | **Indicator** | **Location** | **RH1** | **NE** | **SO** | **NW** |
| --- | --- | --- | --- | --- | --- | --- |
| Magnesium | N1 | NE | 0.209 | - | - | - |
|  |  | SO | 0.597 | 0.017 | - | - |
|  |  | NW | 0.830 | 0.042 | 0.562 | - |
|  |  | WWTP | 0.015 | 0.000 | 0.015 | 0.002 |
|  | N2 | NE | 0.571 | - | - | - |
|  |  | SO | 0.790 | 0.043 | - | - |
|  |  | NW | 0.790 | 0.034 | 0.903 | - |
|  |  | WWTP | 0.002 | 0.000 | 0.000 | 0.000 |
| Potassium | N1 | NE | 0.160 | - | - | - |
|  |  | SO | 0.123 | 0.001 | - | - |
|  |  | NW | 0.770 | 0.048 | 0.016 | - |
|  |  | WWTP | 0.013 | 0.000 | 0.174 | 0.000 |
|  | N2 | NE | 0.423 | - | - | - |
|  |  | SO | 0.227 | 0.008 | - | - |
|  |  | NW | 0.922 | 0.109 | 0.101 | - |
|  |  | WWTP | 0.001 | 0.000 | 0.002 | 0.000 |
| Sodium | N1 | NE | 0.193 | - | - | - |
|  |  | SO | 1.000 | 0.014 | - | - |
|  |  | NW | 0.314 | 0.324 | 0.051 | - |
|  |  | WWTP | 0.314 | 0.000 | 0.230 | 0.001 |
|  | N2 | NE | 0.418 | - | - | - |
|  |  | SO | 0.973 | 0.110 | - | - |
|  |  | NW | 0.665 | 0.665 | 0.207 | - |
|  |  | WWTP | 0.041 | 0.000 | 0.002 | 0.000 |
| Chloride | N1 | NE | 0.104 | - | - | - |
|  |  | SO | 0.934 | 0.006 | - | - |
|  |  | NW | 0.222 | 0.506 | 0.020 | - |
|  |  | WWTP | 0.286 | 0.000 | 0.124 | 0.000 |
|  | N2 | NE | 0.295 | - | - | - |
|  |  | SO | 0.947 | 0.065 | - | - |
|  |  | NW | 0.424 | 0.777 | 0.101 | - |
|  |  | WWTP | 0.038 | 0.000 | 0.001 | 0.000 |
| Calcium | N1 | NE | 0.012 | - | - | - |
|  |  | SO | 0.293 | 0.000 | - | - |
|  |  | NW | 0.381 | 0.001 | 0.009 | - |
|  |  | WWTP | 0.012 | 0.000 | 0.036 | 0.000 |
|  | N2 | NE | 0.067 | - | - | - |
|  |  | SO | 0.317 | 0.001 | - | - |
|  |  | NW | 0.682 | 0.004 | 0.048 | - |
|  |  | WWTP | 0.001 | 0.000 | 0.000 | 0.000 |
| PMMoV | N1 | NE | 0.209 | - | - | - |
|  |  | SO | 0.597 | 0.017 | - | - |
|  |  | NW | 0.830 | 0.042 | 0.562 | - |
|  |  | WWTP | 0.015 | 0.000 | 0.015 | 0.002 |
|  | N2 | NE | 0.571 | - | - | - |
|  |  | SO | 0.790 | 0.043 | - | - |
|  |  | NW | 0.790 | 0.034 | 0.903 | - |
|  |  | WWTP | 0.002 | 0.000 | 0.000 | 0.000 |
